# Can Catastrophe Theory explain expansion and contagious of Covid-19?

**DOI:** 10.1101/2021.01.02.21249133

**Authors:** Marco Antonio Leonel Caetano

## Abstract

Since SARS-Cov-2 started spreading in China and turned into a pandemic disease called Covid-19, many articles about prediction with mathematical model have appeared in the literature. In addition to models in specialized journals, a significant amount of software was made available, presenting with dashboards spreading of the pandemic for each new. These models are solved by computer simulation of traditional exponential models as a representation of the growth of cases and deaths. Some more accurate models are based on existing variations of SIR model (Susceptible, Infected and Recovered). A third class of study is developed in spatial or probabilistic models as a way of forecasting the effect of Covid-19 around the world. Data on the number of positive cases in all countries, show that SARS-Cov-2 shows great resistance even after strategies of lockdown or social distancing. The purpose of this article is to show how the bifurcation theory, known as Catastrophe Theory, can help to understand why Covid-19 expansion rates change so much and even with low values for a longtime trigger contagion quickly and abruptly.

The Catastrophe Theory was conceived by the mathematician René Thom in the 60s with wide applications in works in the 70s. The outbreak of spruce budworm in Canada revealed a very interesting opportunity to test Catastrophe Theory whose explanation for the phenomenon was widely debated in the academic world. Inspired by the same mathematical approach to this phenomenon in Canada in the 1970s, we applied the Catastrophe Theory in the current Covid-19 pandemic. We observed that sudden outbreaks occur when the carrying capacity and the rate of expansion of the virus reach a region of bifurcation on the cusp surface. With actual Covid-19 data obtained from WHO, we fitted the dynamic model using the particle swarm technique and compared the results in the bifurcation plan with the Covid-19 outbreaks in different regions of the world. It is possible in many cases to observe the trajectory of the parameters between limit points in the bistable region and the consequent explosion of cases observed for each country assessed.

## 1. INTRODUCTION

A new human coronavirus emerged between November and December 2019 in Wuhan, China, and its spread became a pandemic declared by the World Health Organization on 11 March 2020. The spread of the pandemic was rapid and with high mortality rates, leading the WHO to declare the pandemic with a specific name, calling this disease Covid-19. The virus proved to be highly contagious, with waves of infections not dependent on climate, region or demographic characteristics around the world. The virus infects humans when it comes in contact with nose, eyes or mouth and attacks cells in the respiratory tract causing acute respiratory illness syndrome, possibly leading to death.

Even with lockdown in Wuhan and country restrictions on entry of people from China, the expansion of Covid-19 took place quickly and globally. With bigdata technology, numerous websites have provided data and graphs of comparisons between mathematical forecasting models for the spread of the disease, as well as measures of different rates of epidemiology and infectious diseases to alert the world population. (John Hopkins, WHO^(a,b)^). A large production of scientific articles has been made available in several temporary repositories, dealing with the pandemic in several aspects, from biological understanding through epidemiology and infectology, extending to mathematics, physics and computer science (Cotta *et al*, Yang *et al*., Abramov and Junior; Neyens *et al*., Chitanvis, Mlocek e Lew, Abdeljaoued).

We observed, however, that the great majority of articles related to mathematical modeling in the expansion of cases and deaths by Covid-19, used as a focus the forecast or public policies as control in the spread of the pandemic. When we conduct research on articles in mathematical models in specialized journals with a peer-review policy, the same trend is observed, with models and simulations applied to the forecast of cases, deaths or transmissions by Covid-19 (Kucharski *et al*. (2020), Ndairou *et al*. (2020), Ivorra(2020), Liang(2020), Cuevas(2020), Postnikov(2020), Kang *et al*. (2020), Chintalapudi *et al*. (2020), Yousefpour *et al*. (2020), Liu *et al*. (2020), Giordano(2020)).

The mathematical models for forecasting the Covid-19 pandemic outbreak are either based on the well-known model (SIR), or derived from it with some changes in terms of the computational approach widely used in recent dashboards (Anderson and May (1979), Anderson and May (1985), Anderson and May (1992), Bjørnstad *et al*. (2020)).

Observing at data from different countries for positive cases at the beginning of the outbreak in each country, it was possible to note that in all cases SARS-Cov-2 has a slow onset, with some peaks of initial contamination. After a few days with low or moderate infection, the cases increase abruptly. From levels of around 20 or 30 cases per day, the outbreak increases to 200 cases and then to more than 1000 cases per day, exponentially and oscillatory increasing infection.

Covid-19’s behavior is very similar to AIDS in its onset in the 90s, despite much lower lethality. For instance, May and Anderson (1987) observed the same mechanism in HIV infection when describing the incubation period for AIDS. According to them, the variability in the incubation period and because some people develop AIDS and others do not, could be the result of genetic heterogeneity or specific variable types of HIV. The authors noted that the manifestation of the disease would suggest a hazard function, with the likelihood of the virus manifesting over time assuming the Weibull distribution.

The fluctuation of the viral load at the beginning of HIV infection was also observed in Nowak (Nowak *et al*. (1991) e Nowak *et al*. (1995)). The oscillation of the viral load at the beginning of the infection is observed in real data in these articles. After the HIV latency period and small peaks of action, the viral load increases exponentially and uncontrolled if anti-HIV drugs are not administered. Nowadays AIDS is a controllable disease thanks to the cocktail of new and revolutionary drugs that act at the cellular level.

As can be seen at the beginning of the Covid-19 pandemic in Japan and Singapore (WHO 2020^(b)^) in Figure-1, in the first 68 days since the first positive case, the new daily cases fluctuated but still within a control range. Figure-2 shows data after the first 68 days (indicative arrow in the graph) when the Covid-19 outbreak increased in both countries. From the initial figures below 100 cases in the first days, the number of daily cases exceeded 700 in Japan and 1100 in Singapore around day 80 and 100 respectively. This behavior was observed in all countries of the world, from the most populous to the least demographic, from the richest to the poorest.

Nepal (WHO 2020^(b)^) remained with a single case for 61 days, then another 20 days with cases below 10 and then positive cases increased daily. The USA (the country with the highest number of cases and deaths until May / 2020) remained for up to 50 days since the first case with fluctuations and peaks that did not exceed 70 daily cases

**Figure-1.**
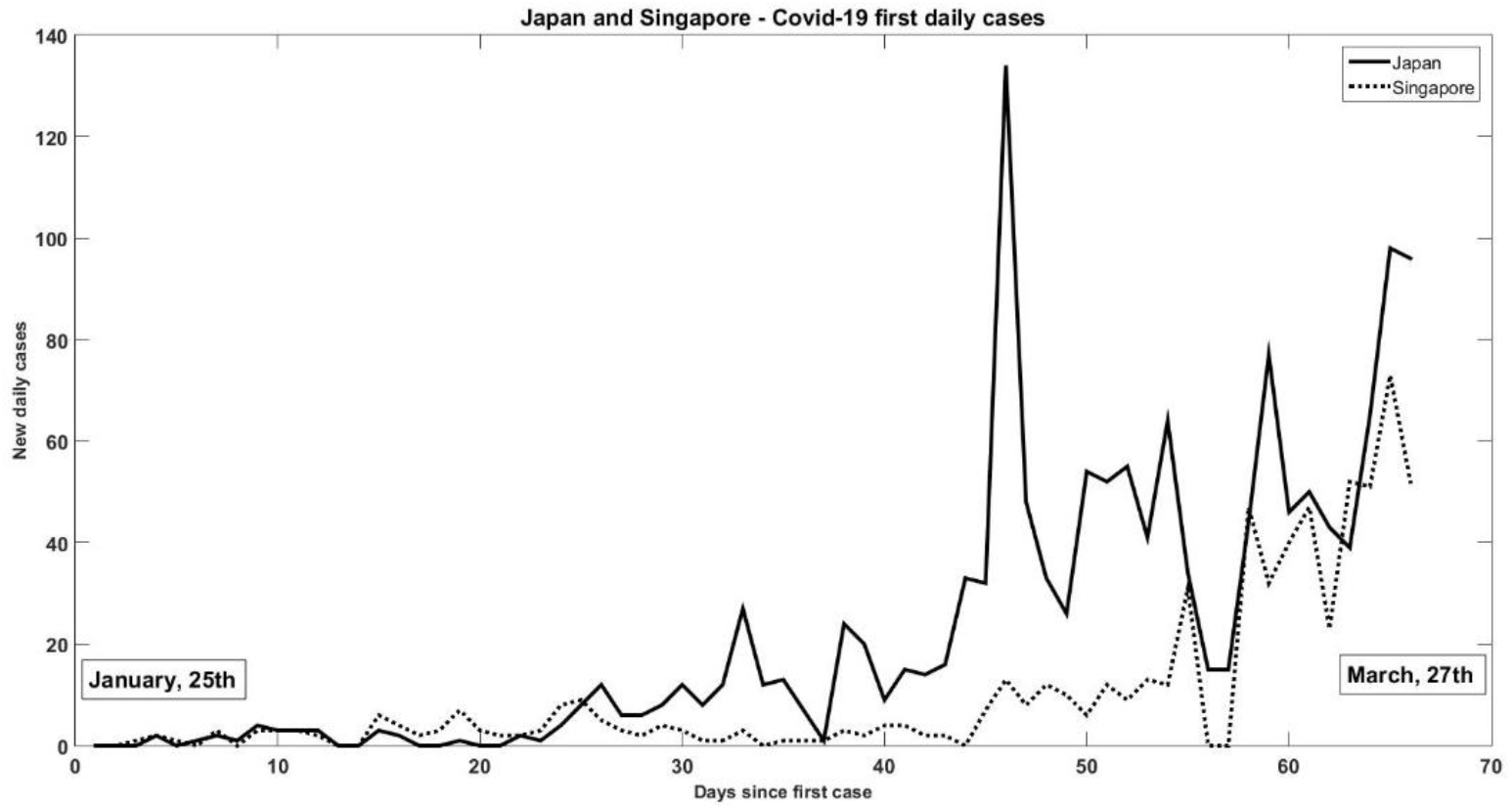
New daily cases in Japan and Singapore.

**Figure-2.**
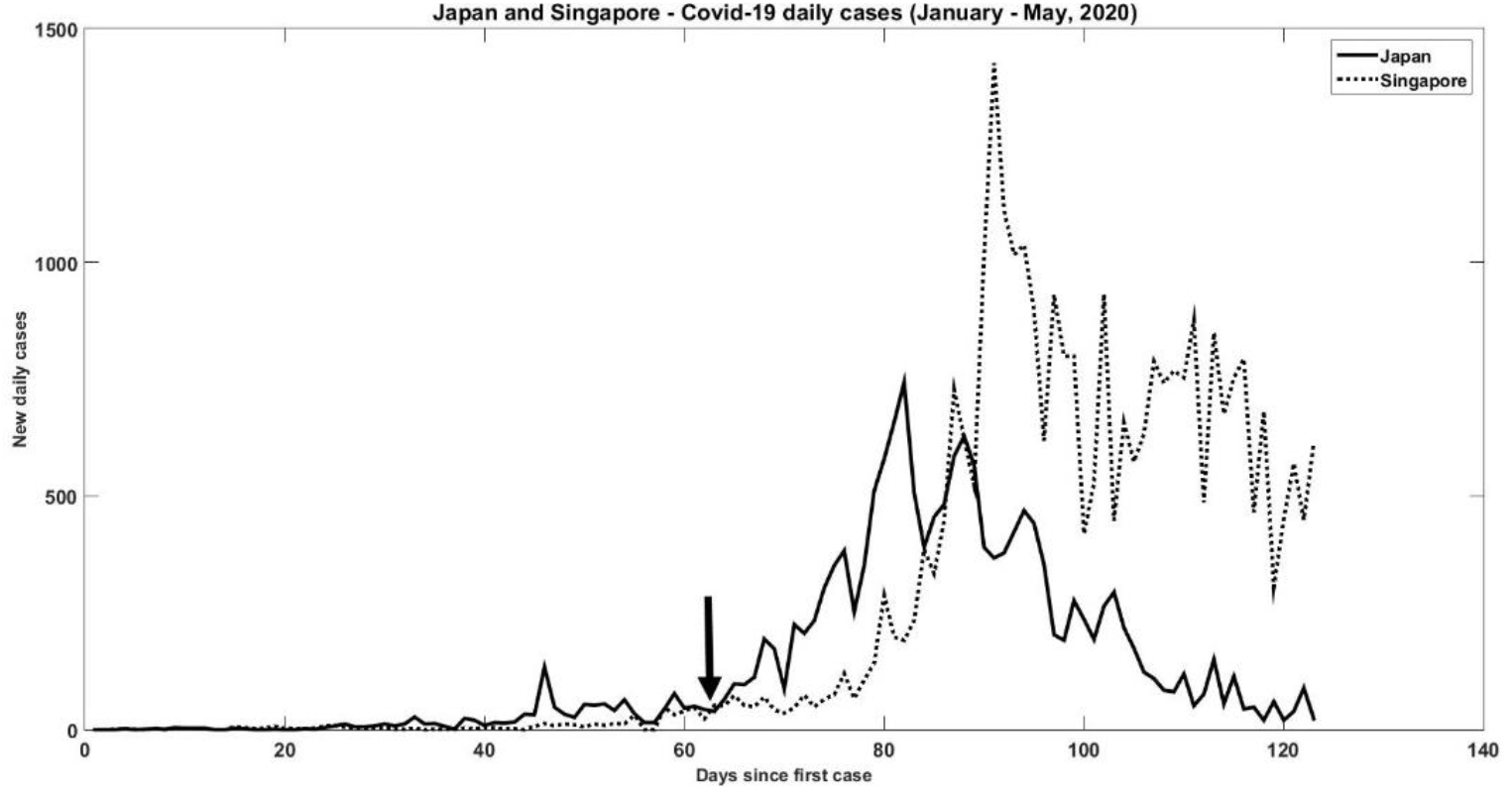
Distribution of daily cases in Japan and Singapore.

The data and behavior of SARS-Cov-2, from the beginning, has its attack and latency behavior very similar to HIV as described before. The equivalence between the two behaviors leads us not only to the oscillation of cases, but oscillation and bifurcation in the values of the parameters of the dynamic system that governs the pandemic. The theory of bifurcation in dynamic systems gave rise to several other important approaches and applications, as in the cases solved for the spruce budworm outbreaks (Ludwig (1978), Casti (1982), Robeva and Murrugarrab (2016)).

After evaluating data from several countries related to Covid-19 caused by SARS-Cov-2, we identified patterns compatible with the oscillations described by the Theory of Bifurcation and the Catastrophe Theory (Zeeman (1977), Woodcock e Davis (1978), Arnold (1986), Gilmore (1993), Castrigiano e Hayes (2004).

The objective of this work is to present how the patterns in the parameter oscillations lead to a sudden increase in the Covid-19 outbreak as predicted by the Catastrophe Theory. After fitting mathematical model in several countries, using the particle swarm algorithm, the work describes how it is possible to understand the explosion of cases of Covid-19 with exponential growth, observing these oscillations in the parameters of the growth rate *r* and carrying capacity *k* for SARS-Cov-2 infection.

## II. METHODS

### A Mathematical Model for Covid-19

In this work we will use the same principle and equations adopted to understand the spruce budworm outbreak. The details about the model and the bifurcation of the main parameters are well known in all literature specializing in ecological models on the outbreak of spruce budworm as in Robeva and Murrugarrab (2016). The authors describe the model and demonstrate how the equations lead to stable and unstable equilibrium conditions, forming bifurcations consistent with the Catastrophe Theory. Adapting the problem for Covid-19, we will represent N(t) as the population of individuals positive for SARS-Cov-2 whose logistic growth model is shown in equation (1).

Assuming as a simplification N≜N(t) the dynamic model adopted in this work will be:

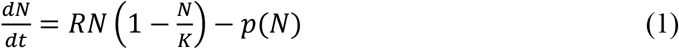

with

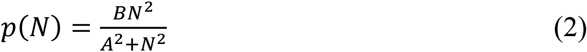

The first term on the right side of the equation (1) indicates that positive cases have logistical growth, but are reduced by the term *p(N)* described in equation (2). In this case *R*: growth rate in the number of infected individuals, *K*: carrying support, *A*: inflection point of the logistic curve e *B*: maximum value of function *p(N)*. Using expansion of the terms in (1) and (2) the complete model is

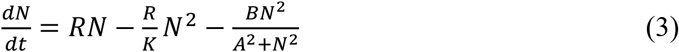

To facilitate the fit of parameters based in actual data from SARS-Cov-2, we will assume that

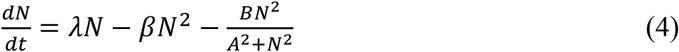

where

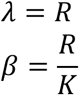

The first two terms on the right side of equation (3) have an analytical solution in the form of an exponential logistic model (considering the third term null)

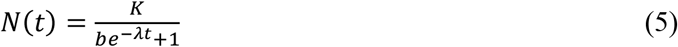

where the parameter b is,

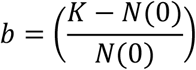

Using the same methodology that are presented in several articles on the bifurcation of parameters for the case of spruce budworm, among them Ludwig (Ludwig *el al*. (1978)), the model (3) becomes interesting to be dimensionless by changing the variable *x* = *N*/*A*. With this change of variable

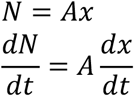

This change leads to equation (3) becoming

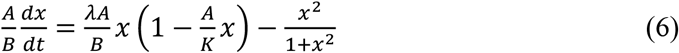

To make equation (5) dimensionless in time

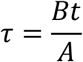

where this change will make equation (1) dimensionless:

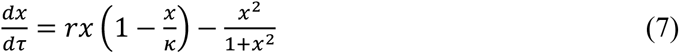

with

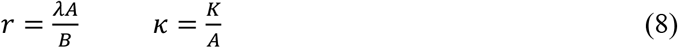

At the equilibrium point *dx/dτ* = 0, then

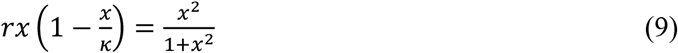

Applying the derivative *x* in equation (8)

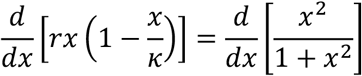

comes the relationship

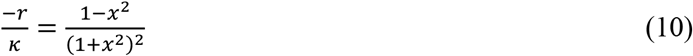

But from equation (8) we have

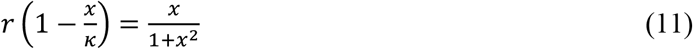

Replacing (10) in (9) is possible find *r* as function of *x*, or,

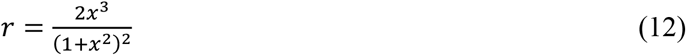

With *r* replacing in (9) is possible find κ,

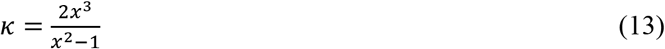

Thus, with the pair of values (κ,*r*) it is possible to evaluate the cases of bifurcation that may lead to the catastrophe predicted in the Catastrophe Theory to assess the sudden increases that occurred in positive cases for covid-19 in all countries.

The methodology for identifying the parameters r e κ for SARS-Cov-2 is:

1. Fit parameters *K*, b and *λ* to equation (5).
2. With K, b and *λ* from step (1), identify A e B from equation (2). Considering the points of p(N) the identification of A and B is easy using, for instance, least squares method, Kalman filter or genetic algorithm.
3. The value of *r* is calculated: 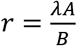
4. The value of *κ* is calculated: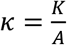

### B. Parameter fit

There are many methods for parameters fitting, from the classic least squares method to more sophisticated methods based on artificial intelligence. When trying to fit the parameters directly for equation (3) the results were not satisfactory, often generating different results for the actual cases. A new strategy was then chosen, which will be explained below in the following algorithm.

Steps to fit:

1. The actual data of positive cases reported to Covid-19 from an initial day to day (or week) D is acquired.
2. Fit parameter to model (5) and make computer simulation to predict distant future.
3. The actual data of positive cases reported to Covid-19 from an initial day to day (or week) D+1 is acquired.
4. Fit parameter to model (5) and make computer simulation to predict distant future.
5. It makes the difference in absolute values between the computer simulation of values obtained from step (2) and step (4). These values are considered points of function p(N) (eq.2).
6. Using p(N) from step(5), insert in abscissa axis the data of the actual positive cases reported up to the day (or week) D + 1 and in ordinate axis the value of the p(N). With *K, b, λ*, A e B calculate *r* e κ from equation (8) building (κ,*r*) for next day (or week) D+1 using (12) and (13).

Figure-3 presents the function p(N) from (2) using difference between predictions D and D+1. Figure-4 shows parameters A e B fit from equation (2). Parameter A is inflexion point of function p(N) and B is maximum value for p(N). One can interpret function p(N) as the braking of Covid-19 in relation to the exponential logistic model of equation (5). This p(N) reduces covid-19 logistical growth and represents the effect of social distancing, lockdown or any other public policy implemented by governments.

**Figure-3.**
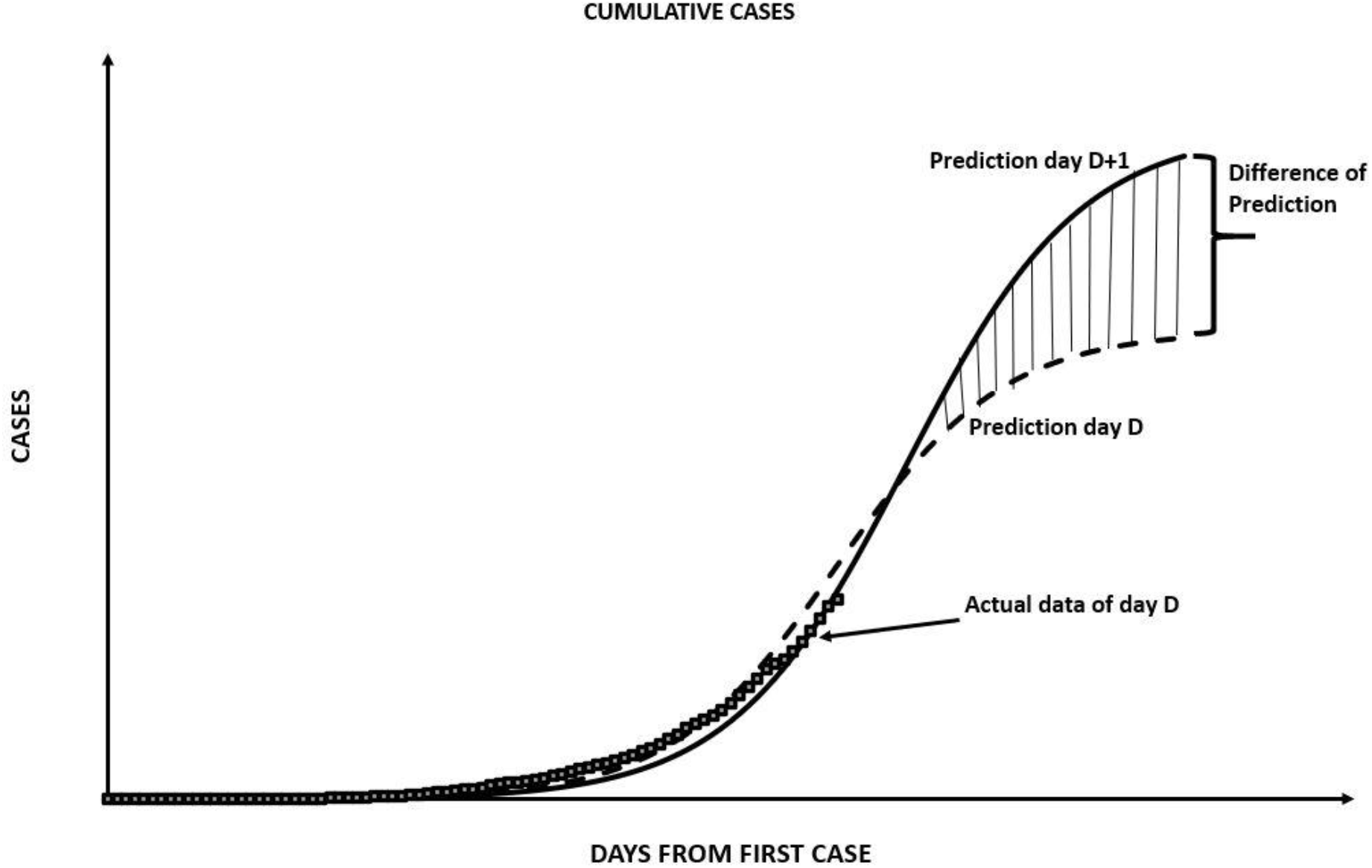
Difference between the forecasts of the logistic model between day (or week) D and day (or week) D + 1.

**Figure-4.**
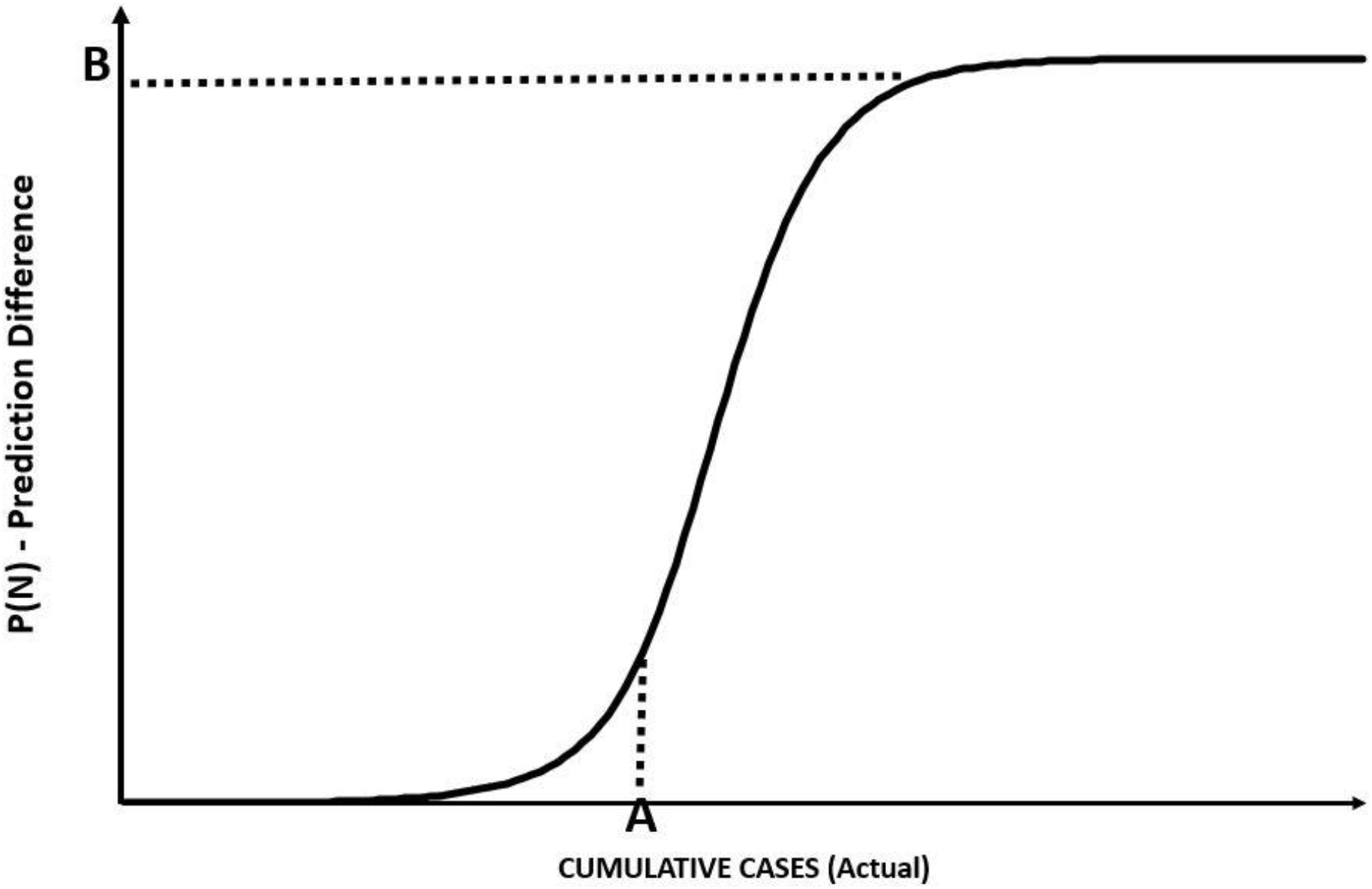
Logistics function p (N) for the difference between D and D + 1 forecasts.

After tests between the classic identification methods and artificial intelligence heuristic models, better results were obtained by using parameter identification according to several articles presented in the literature using the particle swarm algorithm for K, b and λ in equation (5) (Liu (2008), Sendrescu and Roman (2013), Xie *et al*. (2017), Sagar (2017)). In the particle swarm algorithm, the particles were adopted as referring to three different populations, each representing K, b and λ. The initializations of these particles were random with uniform distribution.

The cognition and socialization parameters described in the formulation of the algorithm in the literature, were different to make the convergence better. While cognition was more valued at around 0.05, socialization was 0.005. This allows that when the particles (parameters) converge, they do not disperse as much with irrelevant information from particles further away from the optimum point. The local and global optimums were estimated very high so that the values of the sums of the quadratic differences correct them in the first iteration.

Once the parameters were adjusted (*K*, b and *λ*), steps 1-6 were followed to calculate (κ,*r*). With this pair of parameters, the bifurcation analysis described by Ludwig(1978), Casti (1982) and Robeva and Murrugarrab (2016), is performed according to the scheme shown in Figure-5. The parameters (κ,*r*) are fitted to each new actual data and represented in plane as in Figure-5. The plane is formed by three distinct regions: refuge region (stable), outbreak region (stable region) and the region within the hatched area that has an equilibrium with two stable and one unstable points.

**Figure-5.**
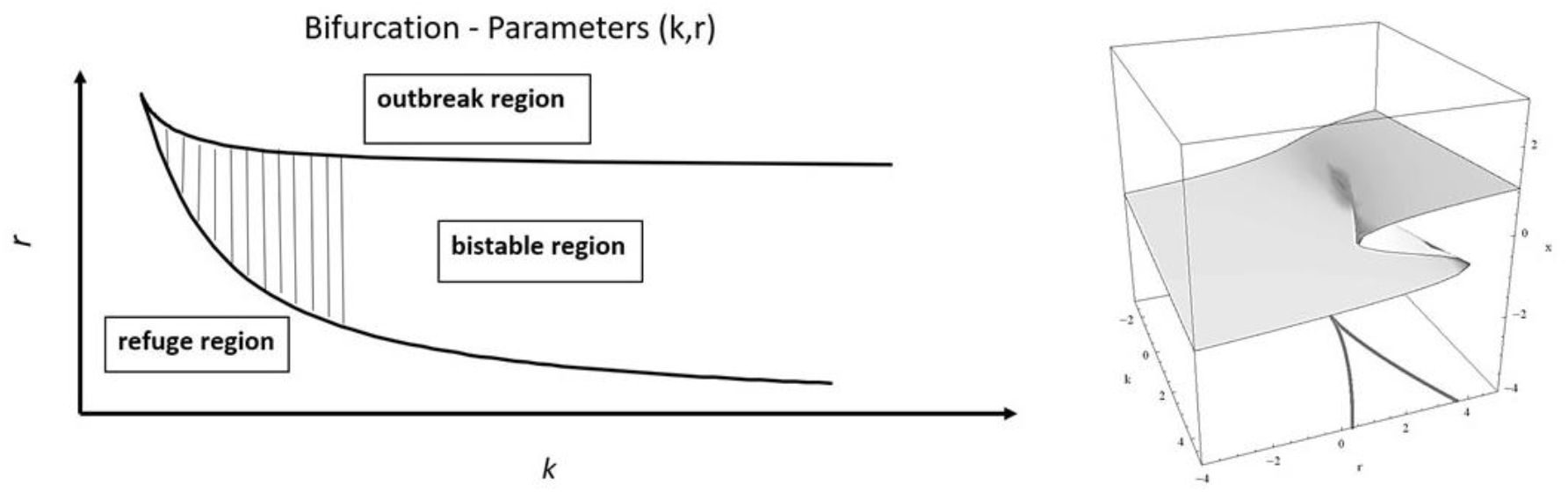
Interpretation of the bifurcation of parameters (κ, r) in 3 different regions.

**Figure-6.**
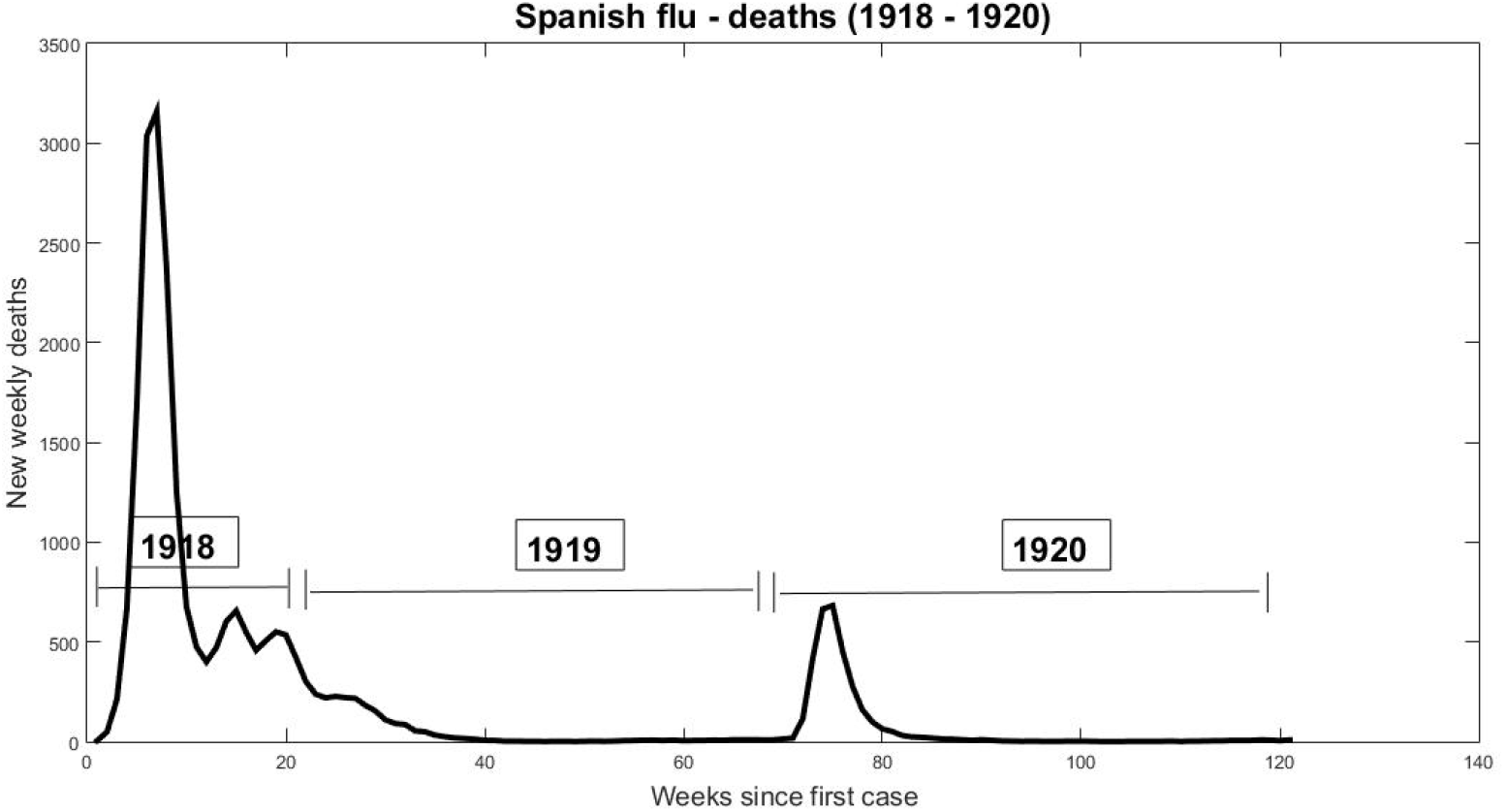
Weekly death data from the Spanish flu (1918-1920)

The two curves that form the regions separated by stable and unstable points are known as the fast path and the slow path of the parameters (κ, r). For values of x starting at 1.0 and traversing the plane with small steps (<< 1.0), we have the upper trajectory of the central curve. For longer steps (>> 1.0), there is the lower curve, forming the “V” region in the center of Figure-5. The value for *r* is calculated from equation (12) and for κ from (13).

According to Casti (1982) these regions form one of the canonical forms of Catastrophe Theory called “cusp catastrophe” (Zeeman (1977), Arnold (1986), Gilmore (1993)). The graph of cup catastrophe is to right in Figure-5. According to Casti(1982) when entering the bistable region the pair (κ, r) jumps abruptly to values that can go to the region of refuge or the region of outbreak, as he verified in the problem of spruce budworm.

### C. The case of the Spanish flu in 1918

As a single example to understand the procedure, data from the Spanish flu pandemic that occurred between 1918 and 1920 were used. The data used in the test were obtained from Collins (1930) for deaths in the 50 largest cities in the United States. Figure-6 presents death data from the 50 largest American cities showing two waves of the pandemic between 1918 and 1919.

Collins data are weekly and were used to fit the model (5) from the previous section, making projections for future weeks from the first week with the first deaths. Figure-7 shows the result for steps 1-4 of the algorithm identification. The dot points are weekly accumulated deaths, the dashed line the projection for week D and the continuous projection line for week D + 1. As an example, the figure presents week D as week 5 while D + 1 is week 6 of the Spanish flu in the USA.

The cumulative death projection curves in Figure-7 were identified with the particle swarm algorithm for each new week of real data. With these data, steps 5 and 6 of the algorithm were applied to calculate the parameter pairs (κ,*r*).

The plane for (κ,*r*) is presented in Figure-8 using data from Table-1 for each week of Spanish flu. The dot point in side left of Figure-8 indicate the trajectory of (κ,*r*) and the corresponding results for weekly deaths in the graph on the right. The data in (1) e (2) are in bistable region in the plane of bifurcation for the weeks 1 and 2. At week 3 there were 213 deaths while at week 4 there were 670 deaths, an increase of 114% while (κ,*r*) went to the region known as the “outbreak” in point (3). Then, since the deaths took the parameters out of the bistable region, the increase of mortality has become out of control.

**Table-1.** Parameters (κ,*r*) identified for the Spanish flu.

| Week | $k$ | $r$ |
| --- | --- | --- |
| 1 | 238 | 0.29 |
| 2 | 30,9 | 0.26 |
| 3 | 57.81 | 1.47 |
| 4 | 350 | 0.53 |
| 5 | 58.4 | 0.09 |
| 6 | 2.35 | 0.51 |

**Table-2.**
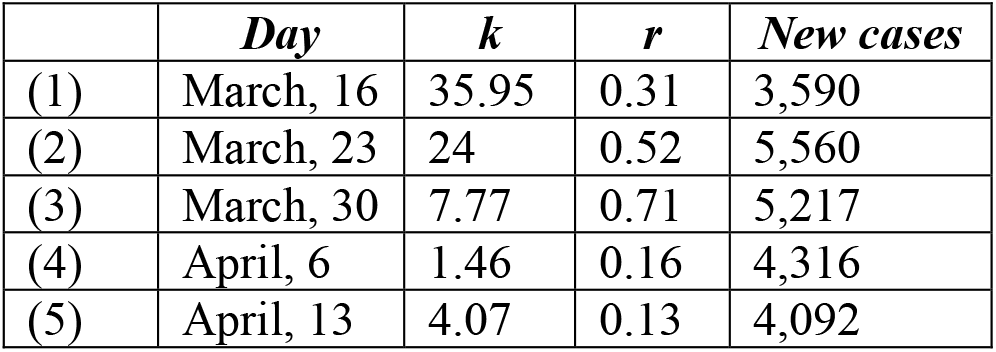
Covid-19 parameters (κ,*r*) identified for Italy.

|  | *Day* | *k* | *r* | *New cases* |
| --- | --- | --- | --- | --- |
| (1) | March, 16 | 35.95 | 0.31 | 3,590 |
| (2) | March, 23 | 24 | 0.52 | 5,560 |
| (3) | March, 30 | 7.77 | 0.71 | 5,217 |
| (4) | April, 6 | 1.46 | 0.16 | 4,316 |
| (5) | April, 13 | 4.07 | 0.13 | 4,092 |

**Figure-7.**
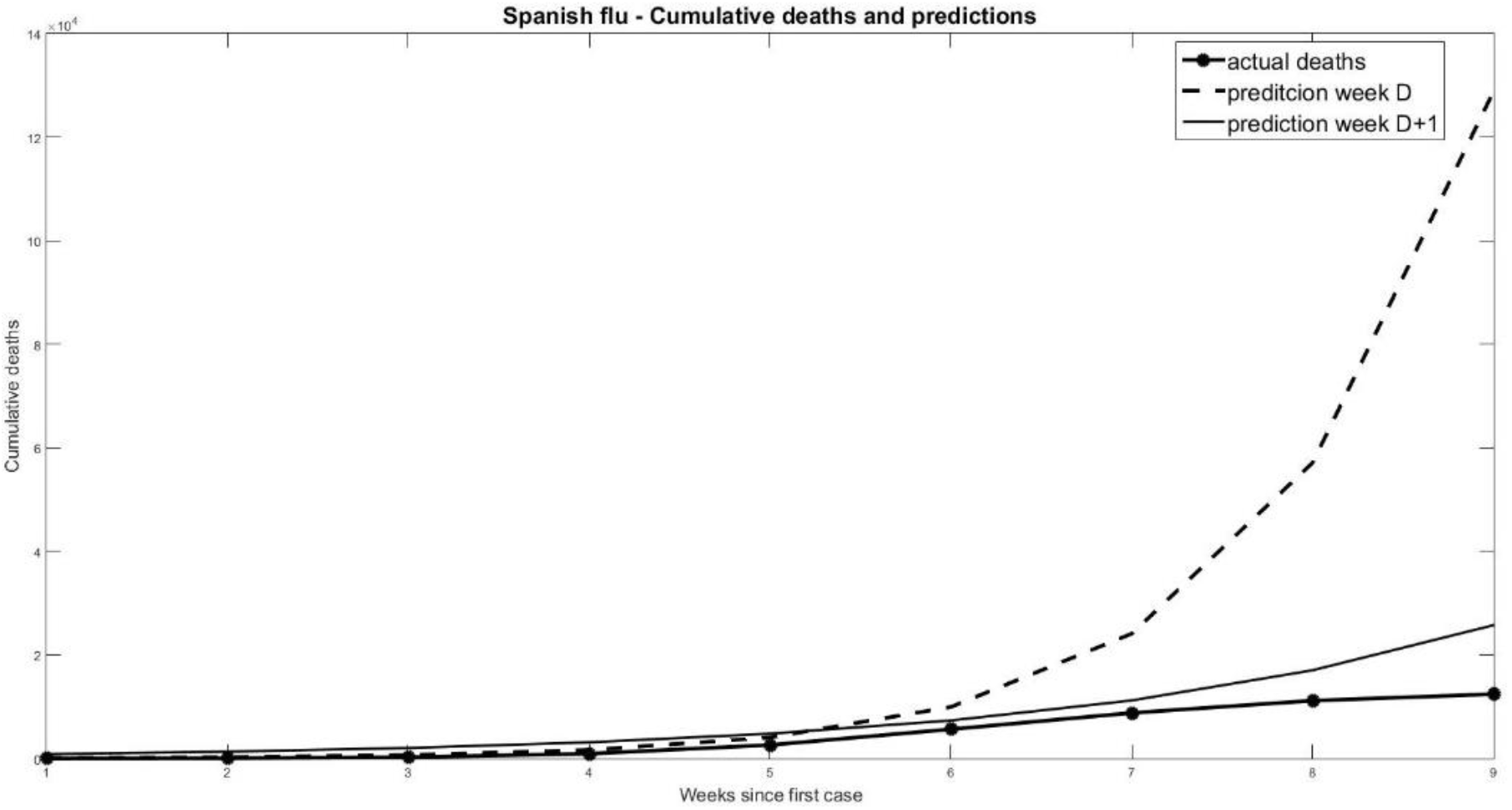
Accumulated weekly deaths from Spanish flu and projections between week 5 and week 6 since infection.

**Figure-8.**
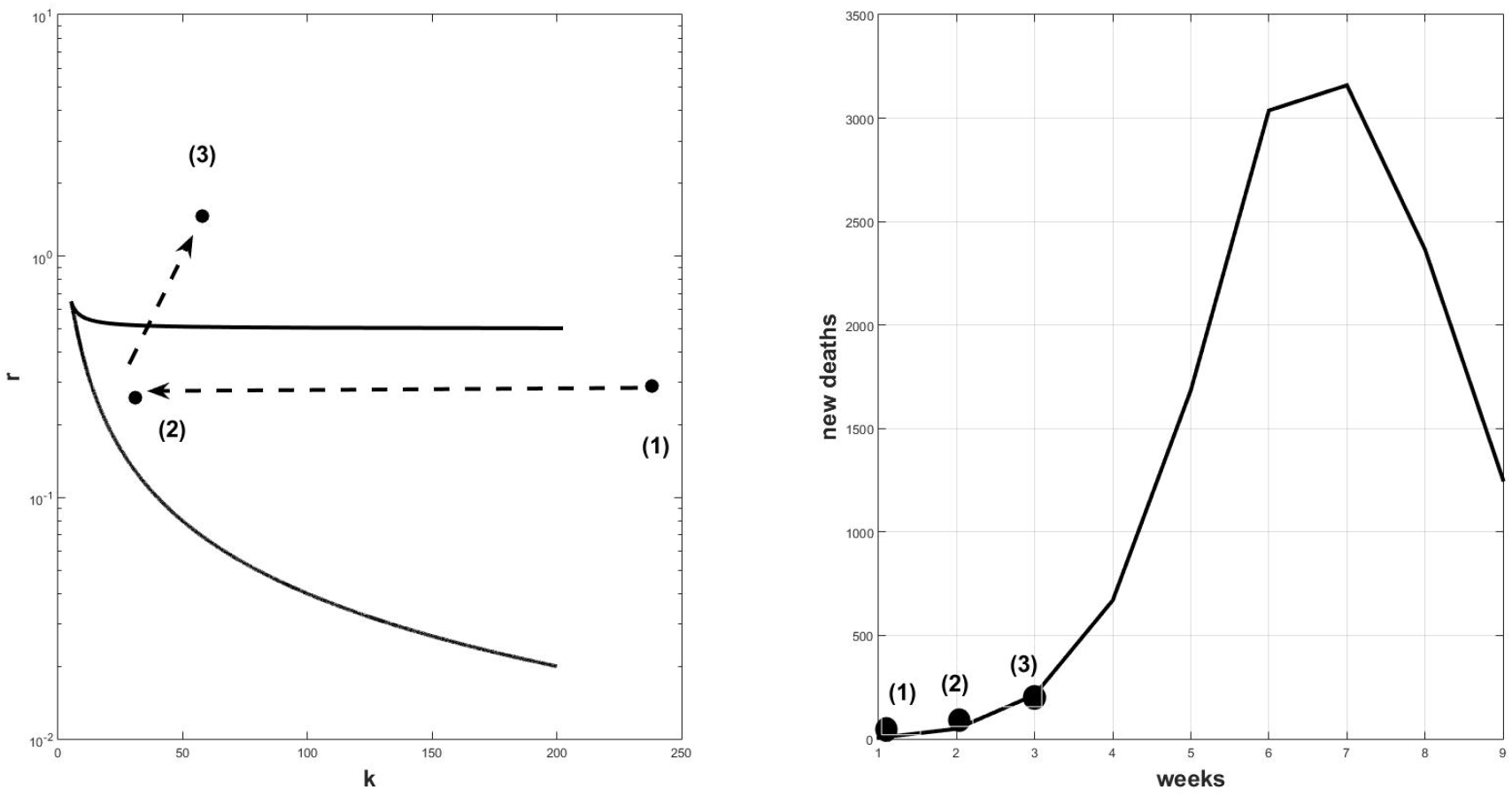
(κ,*r*) bifurcation plan compared to weekly death data for the Spanish flu (1918-1920).

This abrupt and rapid increase is described in Catastrophe Theory and its explanation is due to the fact that when passing in this region, the parameters may fall at the unstable point located within the plane (κ,*r*). This unstable point may cause a significant increase towards the expansion of an outbreak or a sharp drop towards its end.

From the point of view of the Catastrophe Theory it is as if the parameters are passing through the canonical form known as cusp presented in the graph to the right of Figure-5. When entering the bistable region, the parameters may jump to the top fold of the cusp, or they may drop abruptly to the part below, heading towards the refuge region. A combination of the two stable and unstable points within the bistable region will indicate future results in terms of cases or deaths over time.

## III. RESULTS

The methodology presented will be applied to several countries analyzing the expansion of accumulated daily cases reported for the Covid-19 pandemic. The accumulated cases will be used to project the trajectory of (κ,*r*) because the fit of parameter of model (5) is better with few fluctuations and comparisons will be made on the effects in terms of new daily cases. We will observe what happened in each country for different situations when the parameters (κ,*r*) passed through the bistable region in the bifurcation plane.

### A. Italy 2020

After having China as the initial focus of the Covid-19 pandemic, SARS-Cov-2 arrived in Italy causing a rapid expansion in the number of infected people with increases in daily deaths. The first two cases reported in the WHO “situation report” took place on 31 / Jan / 2020 (WHO^(b)^, 2020) arranged with all the data in Figure-9 until April / 2020. The points marked in Figure-9 represent the estimates calculated for the parameters (κ,*r*) in the bifurcation plane.

Table-2 indicates the results obtained using the identification algorithm and the steps presented in the previous section. On March 16, the parameters (κ,*r*) are within the bistable region, indicating a possible sudden increase in the number of positive cases for SARS-Cov-2 in Italy in the following days. The number of cases recorded on that day (cases in last 24 h) was 3,590 while the number of cumulative cases was 24,747 cases.

The parameters in Table-2 were placed on the bifurcation graph to follow the results in the number of daily cases when they left the bistable region. Figure-10 represents the displacement (κ,*r*) for each new day observed. The estimation interval was one week between the data observed for the number of accumulated cases, following the WHO standard. To observe the effects of displacement on the bifurcation plane, we compared the events with Figure 9, which represents the new daily cases.

**Figure-9.**
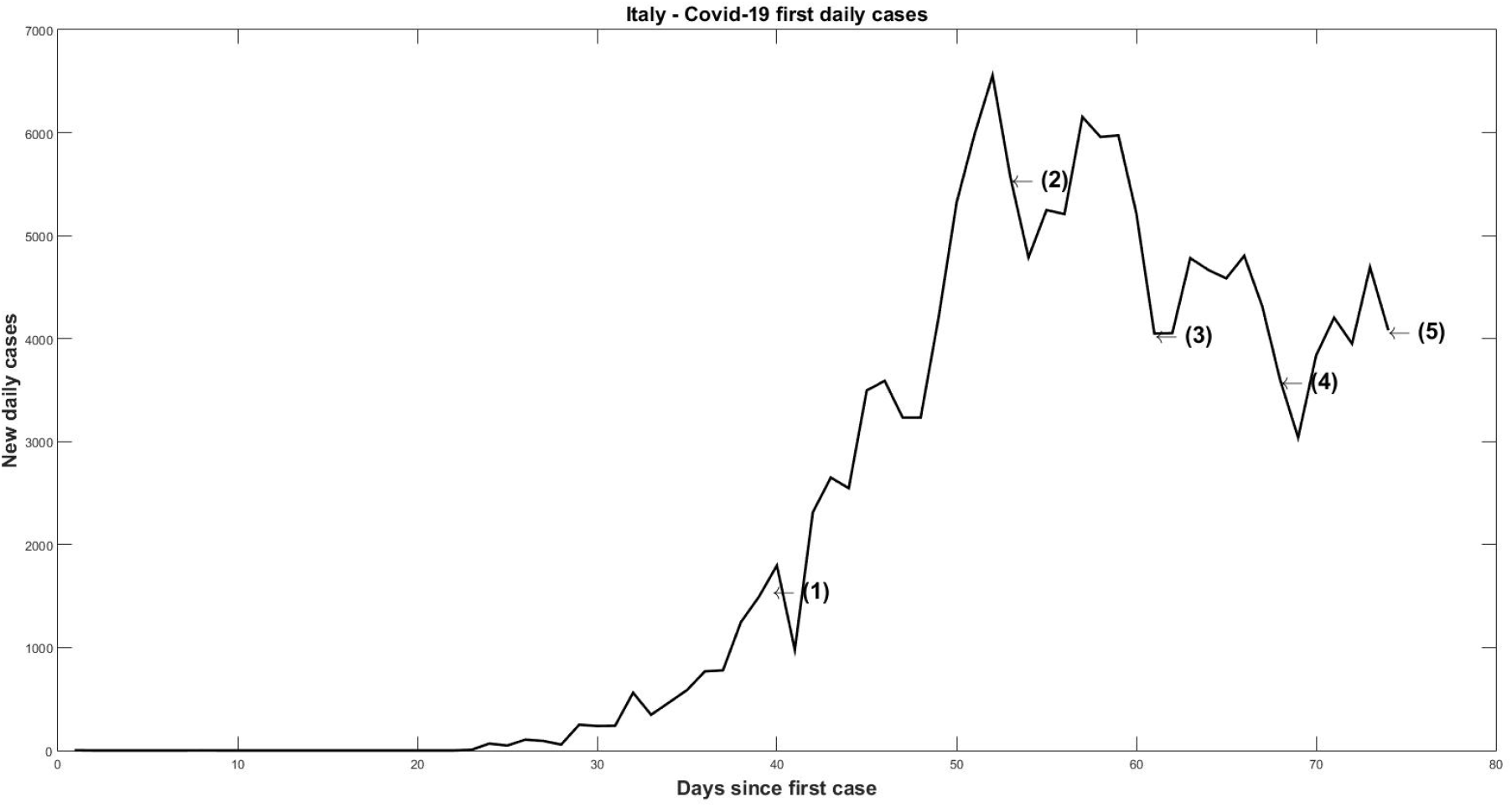
New daily cases in Spain (January - April 2020)

It can be seen in Figure-10 the movement from (1) to (2) towards leaving the bistable region in the bifurcation plane. In the same comparison with Figure-9, we observed a significant increase in cases, going from 3,590 to 5,560 on March 23. In one week new cases increased 54.8%. The number of cases continued to rise a week later with 5,217 cases, but began to decline slowly. The fall was not rapid and it can be seen that from point (3) to points (4) and (5), following the cusp curve of the Catastrophe Theory, the movement did not pass through the bistable region and the points (κ,*r*) followed the path out of the region of abrupt change in the cases.

Figure-11 represents movement of (κ,*r*) from the point of view of the “cusp” catastrophe. The data for (1) with new daily cases jump from the plane of the surface to the top in (2), slowly moving along the upper part of the path up to (3), falling slowly to points (4) and (5).

Beginning on March 13^th^, due to the lockdown and social distancing, the number of new daily cases begins to drop at the same time as deaths. The parameters (κ,*r*) do not return to the bistable region, remaining close to the refuge region. The action of social distancing made the parameters remain outside the bistable region, but a trajectory of (κ,*r*) into that region is a strong indication of loss of control or sudden abandonment of the policy adopted against the pandemic.

### B. South Korea 2020

South Korea was one of the successful cases in combating Covid-19, massively testing its inhabitants and maintaining a strong policy of social distancing. Of the 813 officially registered cases on February 29, there were only 30 cases on April 11, as can be seen in Figure 12. In comparison with the case of Italy, we took points in the bifurcation plan for the controlled phase of the disease, indicated by numbers (1) to (6) in the figure. The idea is to observe whether these controlled cases maintain the parameters (κ,*r*) in the refuge region and how the movement would be correlated with the real data of the new cases.

**Figure-10.**
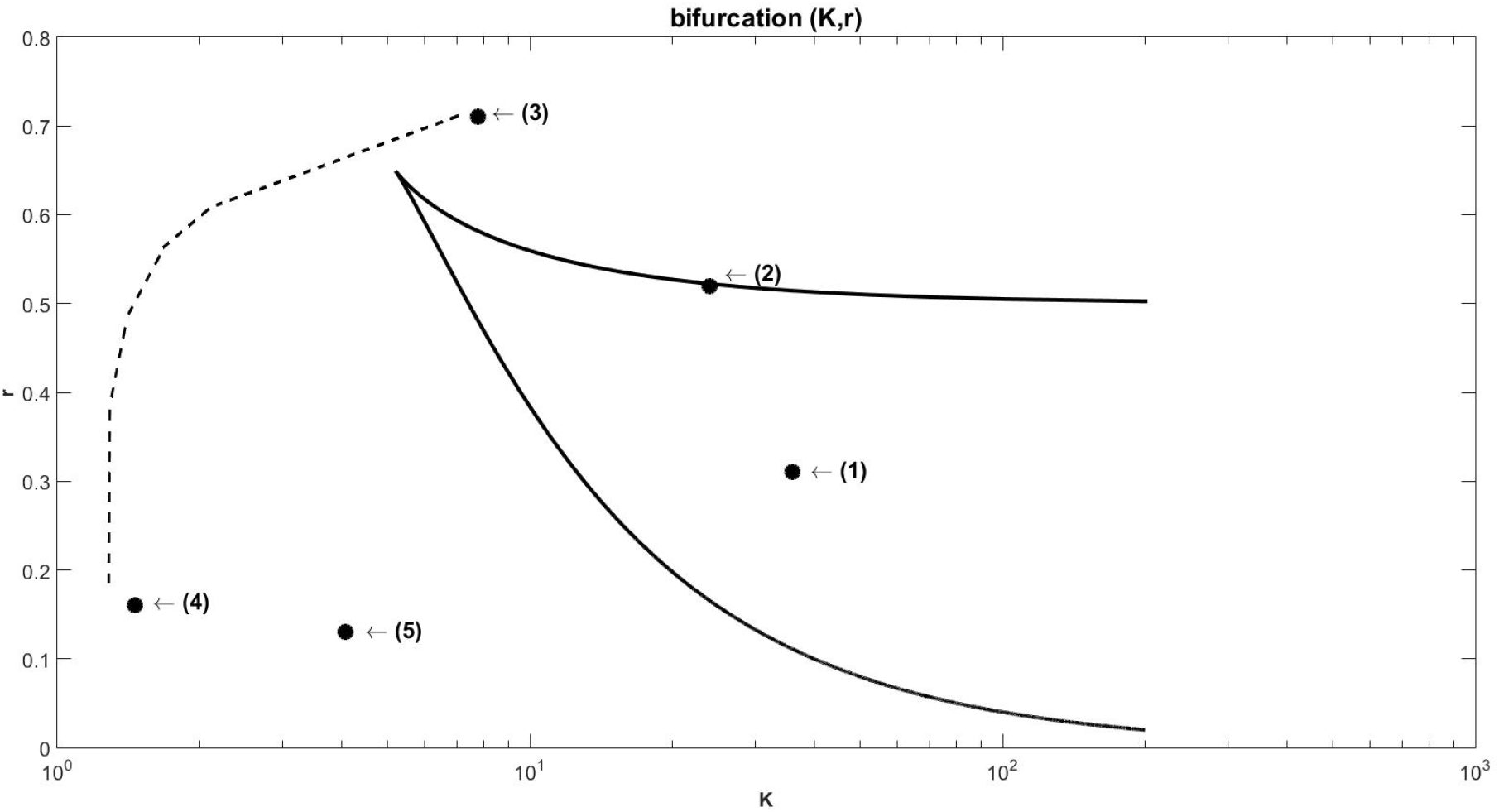
Bifurcation plan with parameters (κ,*r*) estimated for Italy.

**Figure-11.**
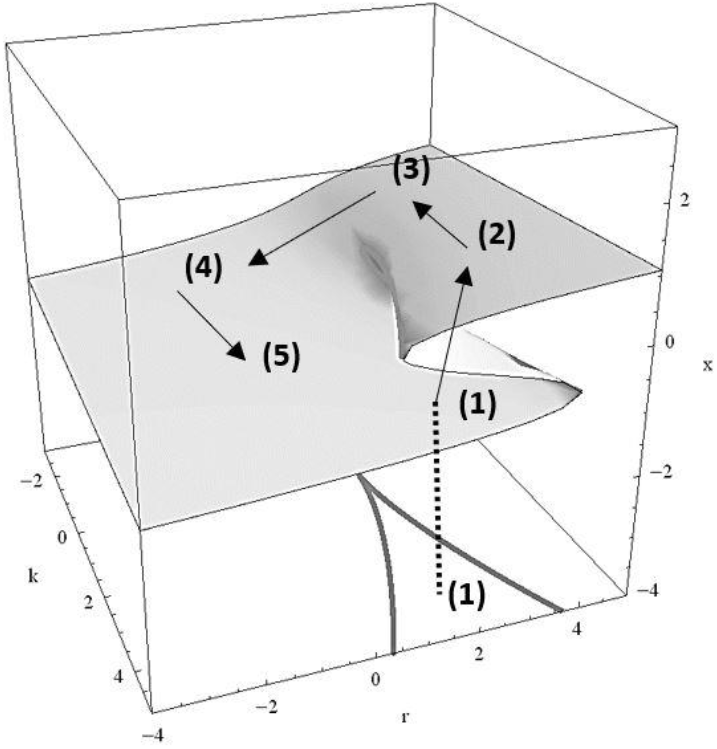
Representation of the movement of the parameters (κ,*r*) in the “cusp” catastrophe with data from Italy.

**Figure-12.**
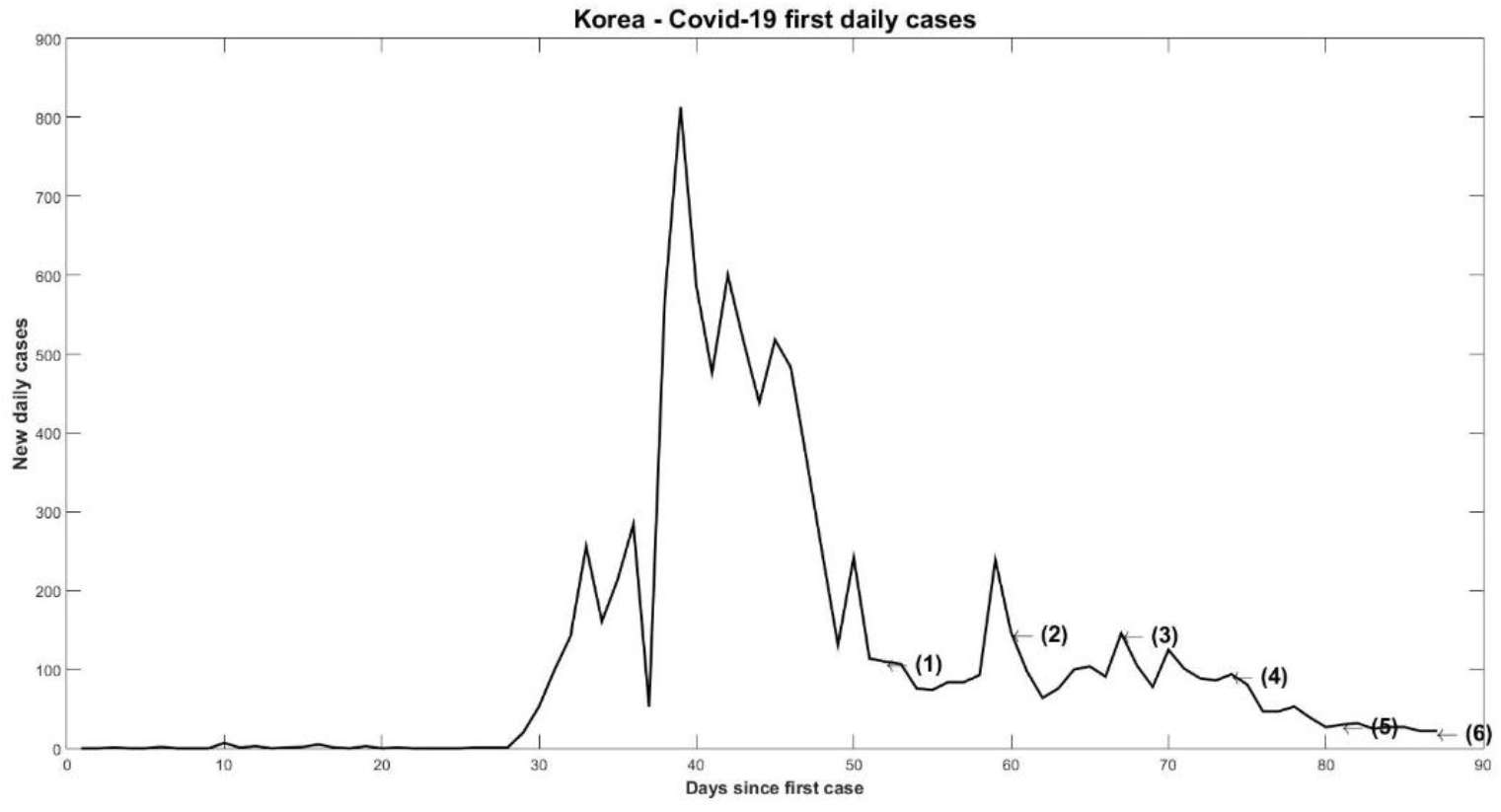
Daily new cases in Korea (January - April 2020)

Table-3 lists the numbers in Figure-13 with the parameters (κ,*r*) identified with Korean data for Covid-19. In particular, we observed a sharp drop between April 4 and April 11, with a reduction of 68%. Of the 94 daily cases in the following week, only 30 new cases were registered. The identification of (κ,*r*) was performed every 7 days so that the changes in the accumulated case curves were significant, since they are the accumulated cases used for the projection and identification, as explained in Figure-3 on the algorithm.

**Table-3.** Covid-19 (κ, r) parameters identified for Korea.

|  | *Day* | *k* | *r* | *New cases* |
| --- | --- | --- | --- | --- |
| (1) | March, 13 | 1.48 | 0.124 | 110 |
| (2) | March, 21 | 2.19 | 0.68 | 147 |
| (3) | March, 28 | 15.32 | 0.098 | 146 |
| (4) | April, 4 | 15.90 | 0.32 | 94 |
| (5) | April, 11 | 3.04 | 0.67 | 30 |
| (6) | April, 17 | 316.3 | 0.01 | 22 |

**Table-4.** Covid-19 (κ, r) parameters identified for Brazil.

|  | *Day* | *k* | *r* | *New cases* |
| --- | --- | --- | --- | --- |
| (1) | April, 22 | 39.90 | 0.147 | 2,498 |
| (2) | April, 25 | 66.00 | 0.520 | 3,503 |
| (3) | April, 28 | 10.40 | 0.860 | 4,613 |
| (4) | May, 1 | 12.83 | 0.080 | 7,218 |
| (5) | May, 7 | 219.13 | 0.036 | 10,381 |
| (6) | May, 8 | 17.94 | 0.071 | 10,010 |
| (7) | May, 11 | 6.93 | 0.395 | 6,760 |

The bifurcation plan for the case of Korea can be seen in Figure-13, including the bistable region. We can observe in particular points (4) and (5) with the trajectory indicating the exit of the bistable region from point (4) to point (5). It was not possible to accurately identify the trajectory between these points, since the values are close when compared in terms of accumulated data (from 94 new cases to 30 new and 10156 cases accumulated on April 4 to 10480 on April 11). But due to the significant drop of 68%, perhaps the trajectory had an abrupt drop in the lower part of the bistable region at point (4) following towards point (5).

**Figure-13.**
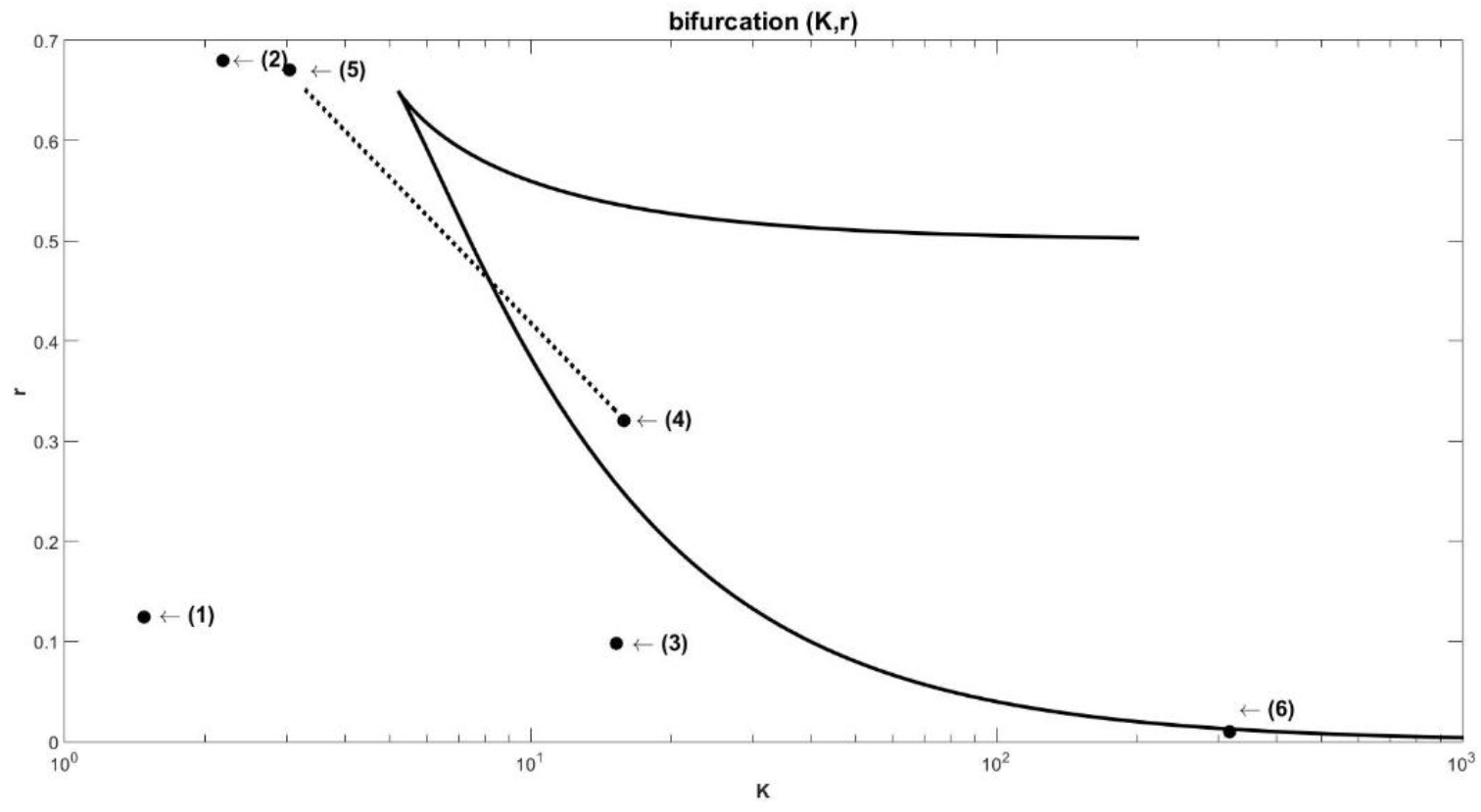
Bifurcation plan with estimated parameters (κ,*r*) for Korea.

**Figure-14.**
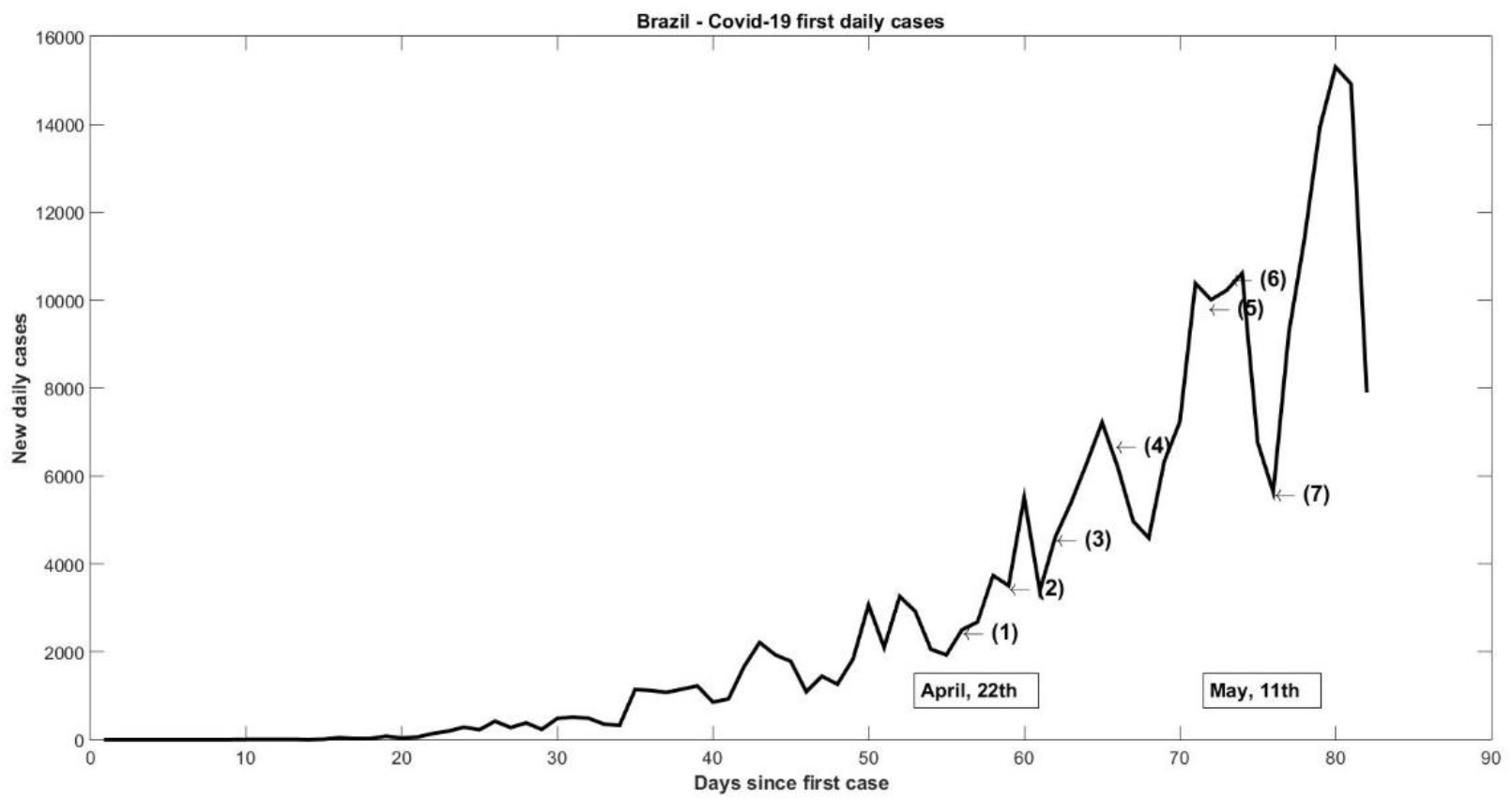
New daily cases in Brazil (April - May 2020)

As in those days the pandemic was already in the control phase in Korea, even with data sampled every 7 days, the identification loses precision regarding the location of the parameters (κ,*r*). Identification is best when changes in the accumulated data curve are significant. For the convergence phase of the accumulated cases, the region hatched in Figure-3 is small and fit on the logistic curve becomes difficult.

Despite this, even at the most distant point (6) in the bifurcation plane, it is noticed that in the disease control phase, the parameters (κ,*r*) barely enter the bistable region, showing that this region was known as a refuge really serves as a refuge. a control indicator for the data obtained from Covid-19.

### C. Brazil 2020

Brazil is an emblematic case of uncontrolled Covid-19 that needs a more detailed study. Since the beginning, with the denial of the pandemic by President Jair Bolsonaro and his followers, a great difficulty in convincing the population about the gravity of the situation has settled in the country. Only after the Supreme Court, the highest body of the Brazilian justice system, determined which governors and mayors would have the prerogative to fight the disease, did sanitary measures begin to be imposed on the population.

Despite this, the uncontrolled disease continued for many months, with three replacements by health ministers and widespread political confusion. These data for both accumulated cases and new daily cases are important for analyzing the development of the disease in the country from the point of view of the Catastrophe Theory.

Figure-14 shows the new daily cases for Brazil, indicating with the numbers (1) to (7) days observed in comparison with the parameters identified in Table-4. As can be seen, in the sampling period there were many fluctuations in new cases. One reason for this was the irregular disclosure of the reports on Covid-19 issued by the municipalities. Many cities did not send data on weekends, barring cases that were only sent at the beginning of each week.

Another reason was the countless openings and closings of commerce, pressured by businessmen from each locality, even instructing protests against social distance or lockdown on account of business.

Finally, another reason was the attempt by the government of Jair Bolsonaro to filter data, changing the form of availability of official data, not reporting the number of accumulated cases, only the new cases that emerged every 24 hours. It was necessary for the Supreme Court and the journalism companies to work on their own collection platform, so that the Brazilian government could go back. Even so, what we saw in the following days were always significant differences between official data and parallel data from journalism.

These facts can all be compared to the results in the calculations in the movement of the parameters (κ, r) in and out of the bistable region in the bifurcation plane, shown in Figure-15. From point (1) to point (3) the number of new daily cases went from 2,498 to 4,613, an increase of 84.6% with the departure of the parameters of the bistable region. In the opposite direction, it is possible to see that while in point (5) the number of cases was 10,381, when leaving the bistable region, new cases fell 34% up to point (7). The reduction was 3,621 cases in just 4 days. When the parameters leave the bistable region for the outbreak region, there is an abrupt increase in cases. When (κ, r) leave for the region of refuge, an abrupt drop in the number of new daily cases is observed.

**Figure-15.**
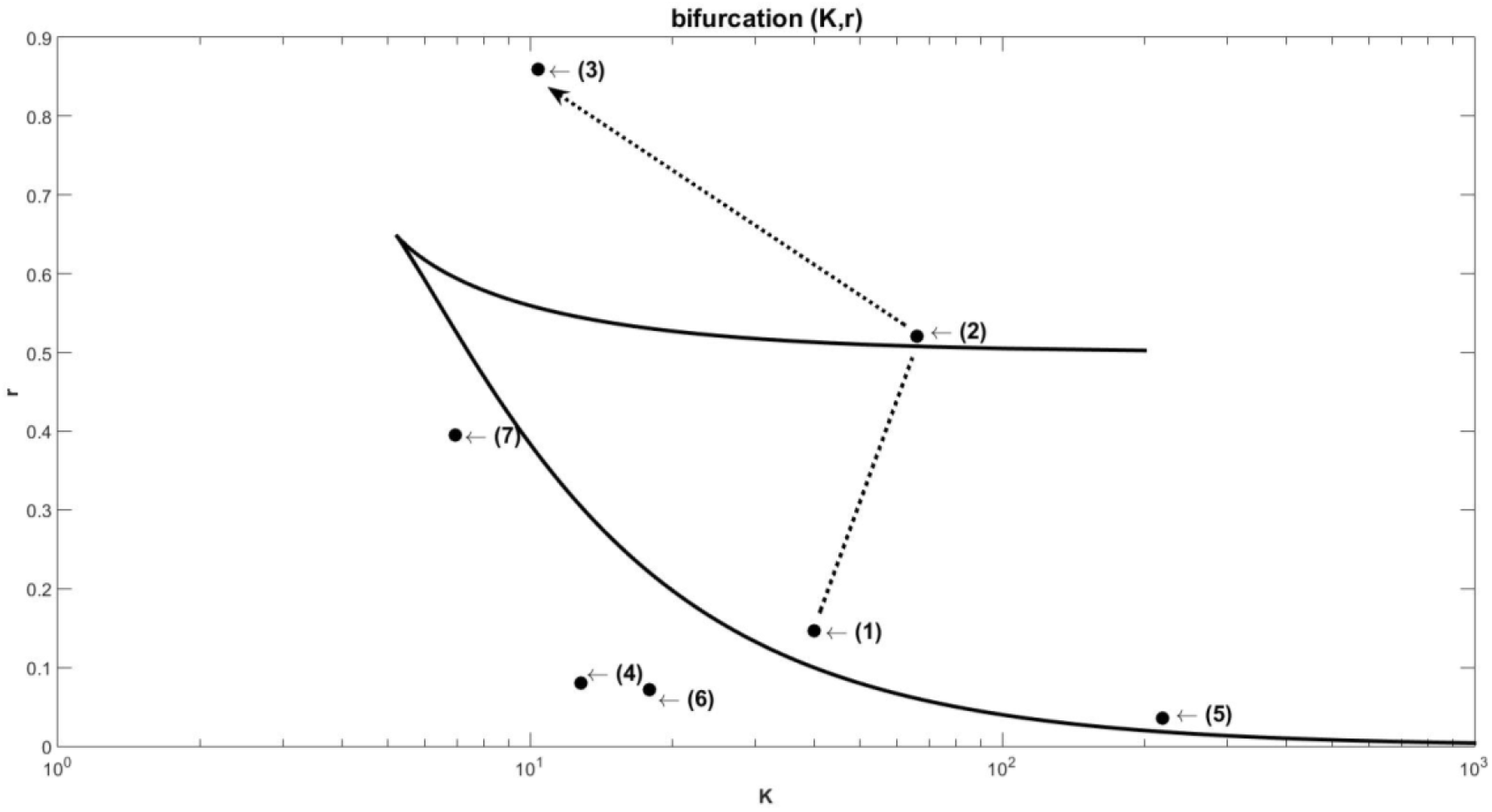
Bifurcation plan with parameters (κ,*r*) estimated for Brazil.

Other movements also cause an increase in cases, but it is not known whether or not they passed through the bistable region because, in some cases, it was not possible to have a good precision due to the proximity in the values of accumulated cases. An example is the movement of (κ, r) from point (3) to point (4) with an increase of 56% in new daily cases, but only 28% in accumulated cases (accumulated cases increased from 66,501 on 28 / April to 85,380 on May 1). From point (3) to point (4) the trajectory may be either outside the bistable region, or the trajectory may have passed the internal limit of that region.

### D. United Kingdom 2020

The British government warned the population about the danger of the new pandemic, but on March 16, it ordered only social distance, asking the population to avoid “non-essential” social contact and unnecessary travel. Stricter measures were adopted on March 23 with mandatory lockdown for three weeks. On March 27, Prime Minister Boris Johnson announced that he was with Covid-19.

Figure-16 shows the new daily cases in the UK, which had a rapid expansion of the pandemic at points (1) to (3) represented in the graph. From point (4) until the end of the sampling for May 9, the cases remained stationed at a high level. After that period, cases started to decline, with the UK going through the relaxation phase of the lockdown.

We identified the parameters (κ, r) following the algorithm presented for all points (1) - (8). Using Table-5, Figure-17 was constructed for the points in the bifurcation plane for the UK.

The same points (1) - (8) noted in Figure-16 are represented in Figure-17 with the parameters (κ,*r*) shown in Table-5. As can be seen, points (1) and (2) represent new daily cases of March 22 and March 28 and they are within the bistable region in the bifurcation plane. When the parameters are in that region, Figure 16 shows an acceleration of the pandemic in the United Kingdom.

**Figure-16.**
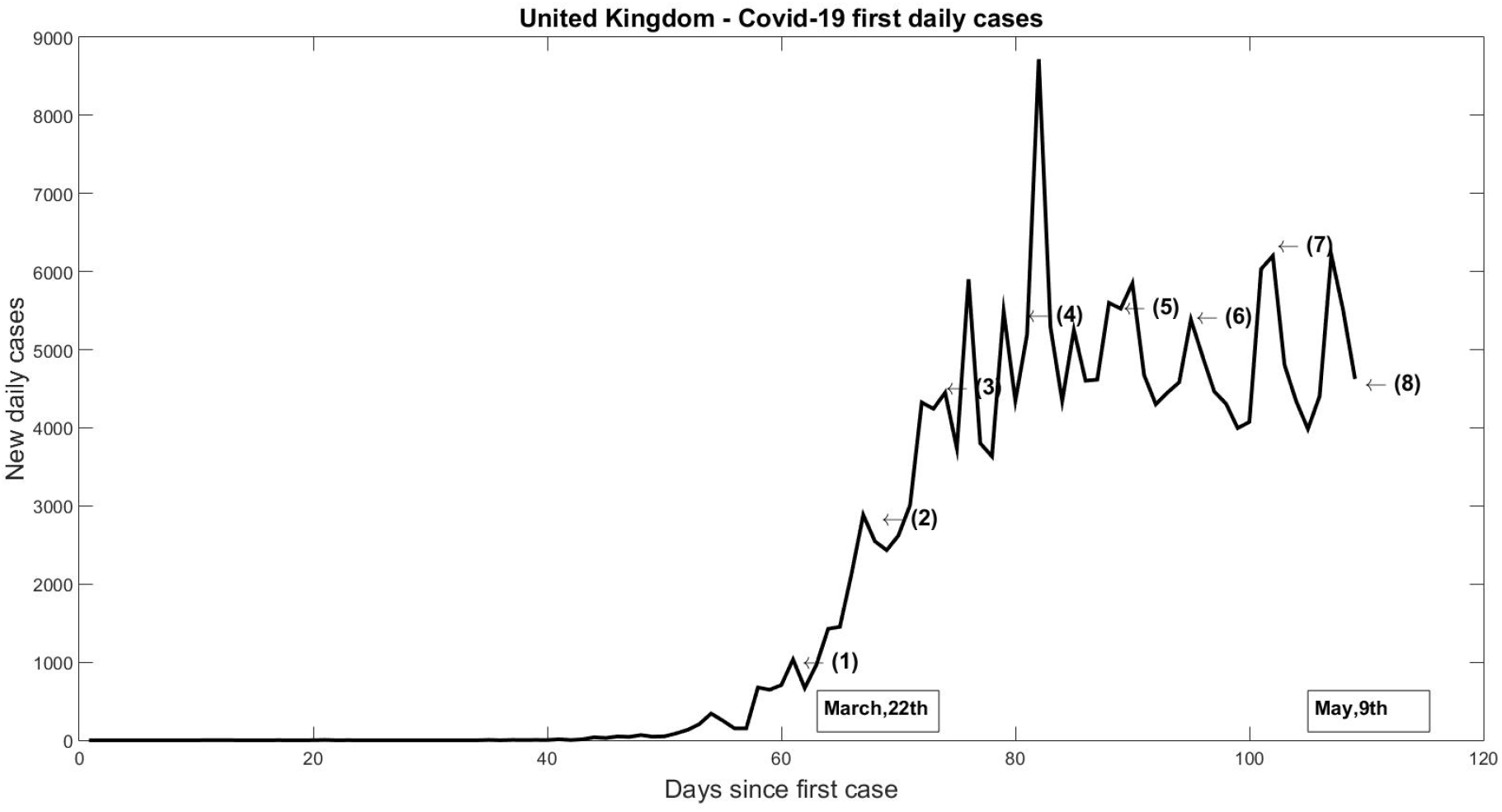
New daily cases in UK (March - May 2020)

**Figure-17.**
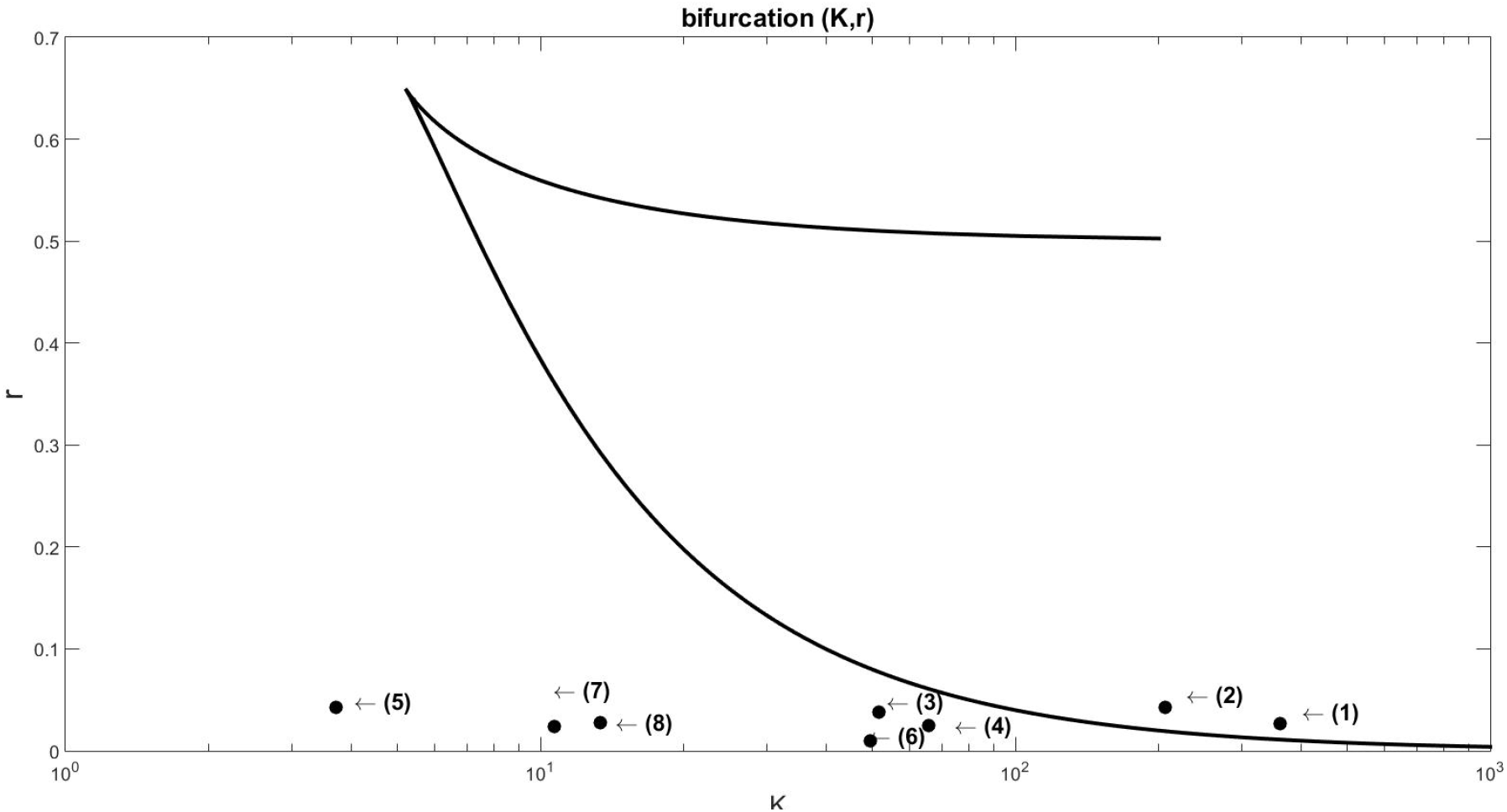
Bifurcation plan with estimated parameters (κ,*r*) for UK.

Until the lockdown decree, the pandemic was out of control in wide expansion with the parameters (κ,*r*) within the bistable region. Point (3) represents the 4th of April, after which the number of accumulated cases continues to increase, but the number of daily cases stabilizes at a high, but controllable level. The points between (4) - (8) move towards the region known as a refuge in Figure-17. The further these parameters become from the bistable region, the more the decline is accentuated in the UK.

Points outside the bistable region and further to the left configure an ideal situation, where the carrying capacity decreases along with the contamination rate. Points increasingly closer to the region of origin of the bifurcation plan show that the pandemic is controlled and outside the region of expansion.

### E. France 2020

The first three cases occurred in France on 24 January, notified to WHO on 25 January. The expansion of Covid-19 was rapid as in all countries, but the control of the pandemic in France was also rapid. On March 16, President Emmanuel Macron announced the lockdown in the country for 15 days, then extended until May 11. The data analyzed in Figure-18 show only the beginning of the expansion of new daily cases. In particular, the points noted as (1) to (4) referring to the 12th, 14th and 16th of March mark the stage of the disease’s most rapid advance. Between points (5) (March 23) to (8) (April 13) the number of accumulated cases continues to increase, but due to the lockdown, new cases oscillate at a high level.

The high volatility in the number of new cases shown in Figure-18 shows that the points are oscillating around some point of equilibrium. After April 13, the curve in Figure 18 shows downward characteristics, compatible with the control and reopening of the French economy. With the accumulated cases, we identified the parameters (κ,*r*) presented in Table-6.

**Table-5.** Covid-19 (κ, r) parameters identified for UK.

|  | *Day* | *k* | *r* | *New cases* |
| --- | --- | --- | --- | --- |
| (1) | March, 22 | 361 | 0.027 | 1,035 |
| (2) | March, 28 | 207 | 0.043 | 2,885 |
| (3) | April, 4 | 51.64 | 0.038 | 4,450 |
| (4) | April, 11 | 65.73 | 0.025 | 5,195 |
| (5) | April, 18 | 3.71 | 0.043 | 5,599 |
| (6) | April, 25 | 49.43 | 0.01 | 5,386 |
| (7) | May, 2 | 10.71 | 0.024 | 6,201 |
| (8) | May, 9 | 13.37 | 0.028 | 4,649 |

**Table-6.** Covid-19 (κ, r) parameters identified for France.

|  | *Day* | *k* | *r* | *New cases* |
| --- | --- | --- | --- | --- |
| (1) | March, 12 | 1149 | 0.03 | 495 |
| (2) | March, 14 | 125 | 0.07 | 780 |
| (3) | March, 16 | 49.21 | 0.15 | 911 |
| (4) | March, 23 | 57.8 | 0.034 | 1,525 |
| (5) | March, 30 | 76.7 | 0.014 | 2,497 |
| (6) | April, 6 | 24.75 | 0.051 | 1,850 |
| (7) | April, 13 | 103.2 | 0.0046 | 1,595 |

**Figure-18.**
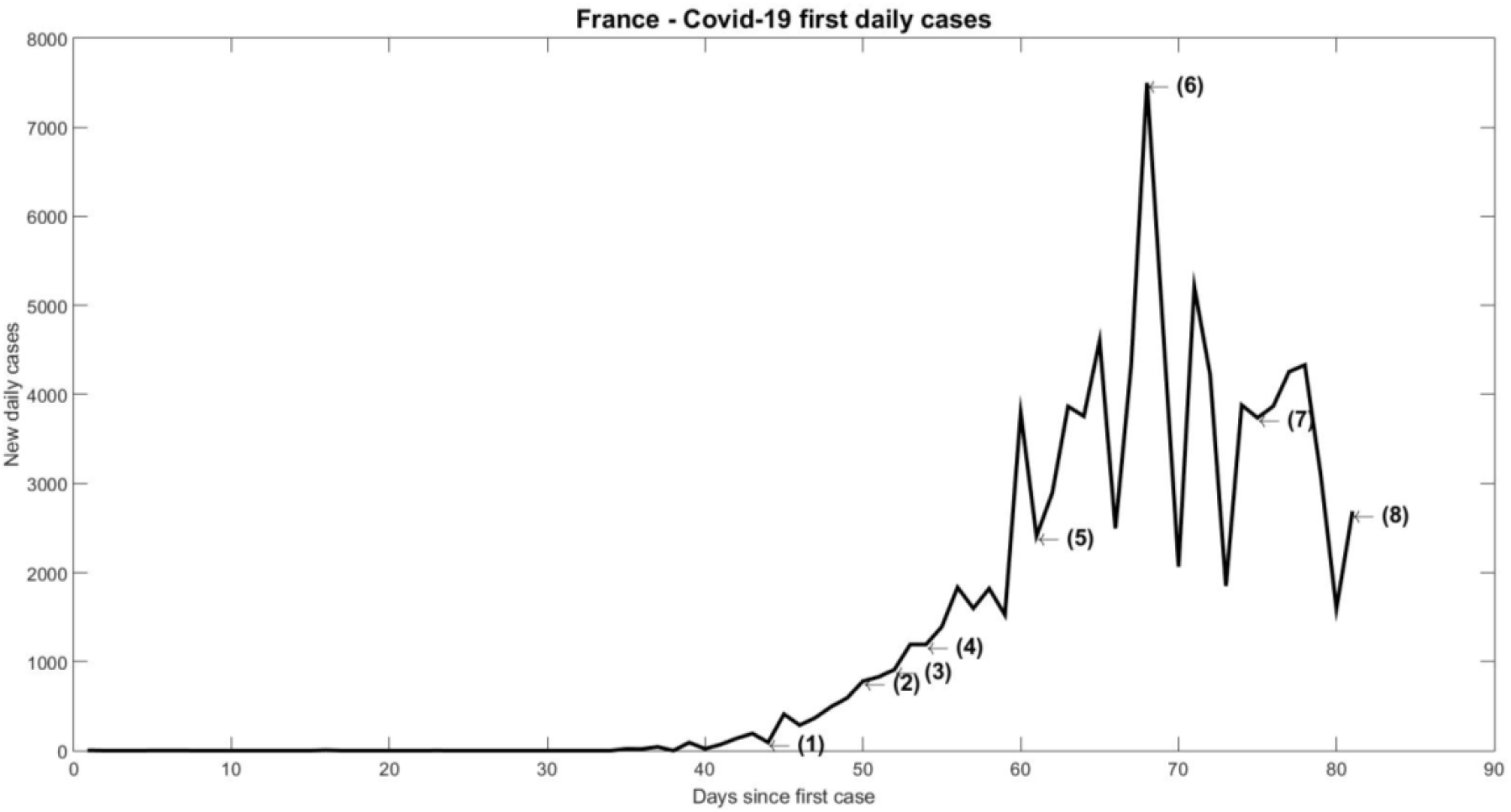
Daily new cases in France (March - April 2020)

As can be seen in Figure-18, the expansion phase of the pandemic was very fast between points (1) to (3). Unlike the other cases, instead of taking data every 7 days, we take between points (1) and (3), case data accumulated every two days. The purpose of this change was to verify if there was a passage of parameters (κ,*r*) in the bistable region. Figure-19 with the data in Table-6 proves that yes, the parameters between March 12 and March 16 pass through the bistable region of the bifurcation plane. It is in this region that rapid expansion is observed when the parameters leave that region at point (3) to point (4).

It is possible to observe in Figure-19 that the beginning of the pandemic in France reflects with the parameters within the bistable region, demonstrating, according to Catastrophe Theory, an abrupt jump when these values migrate out of that region. Points (1) to (3) in this region can be compared with the same points in Figure-18, showing that new cases go from 495 at point (1) to 1,525 at point (4). The increase in the number of new cases was 208% when the parameters (κ,*r*) were from (1) to (4).

**Figure-19.**
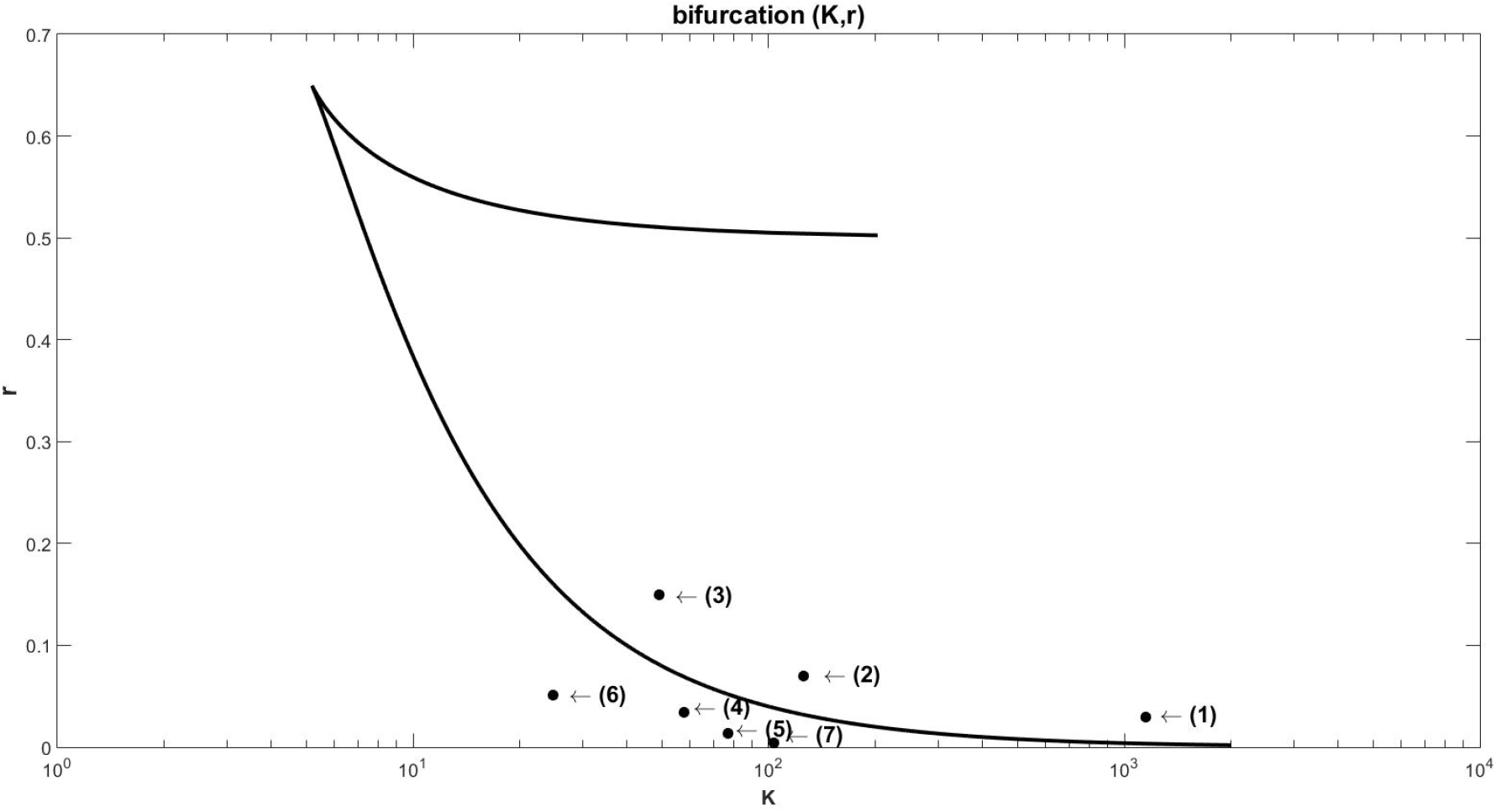
Bifurcation plan with estimated parameters (κ,*r*) for France.

**Figure-20.**
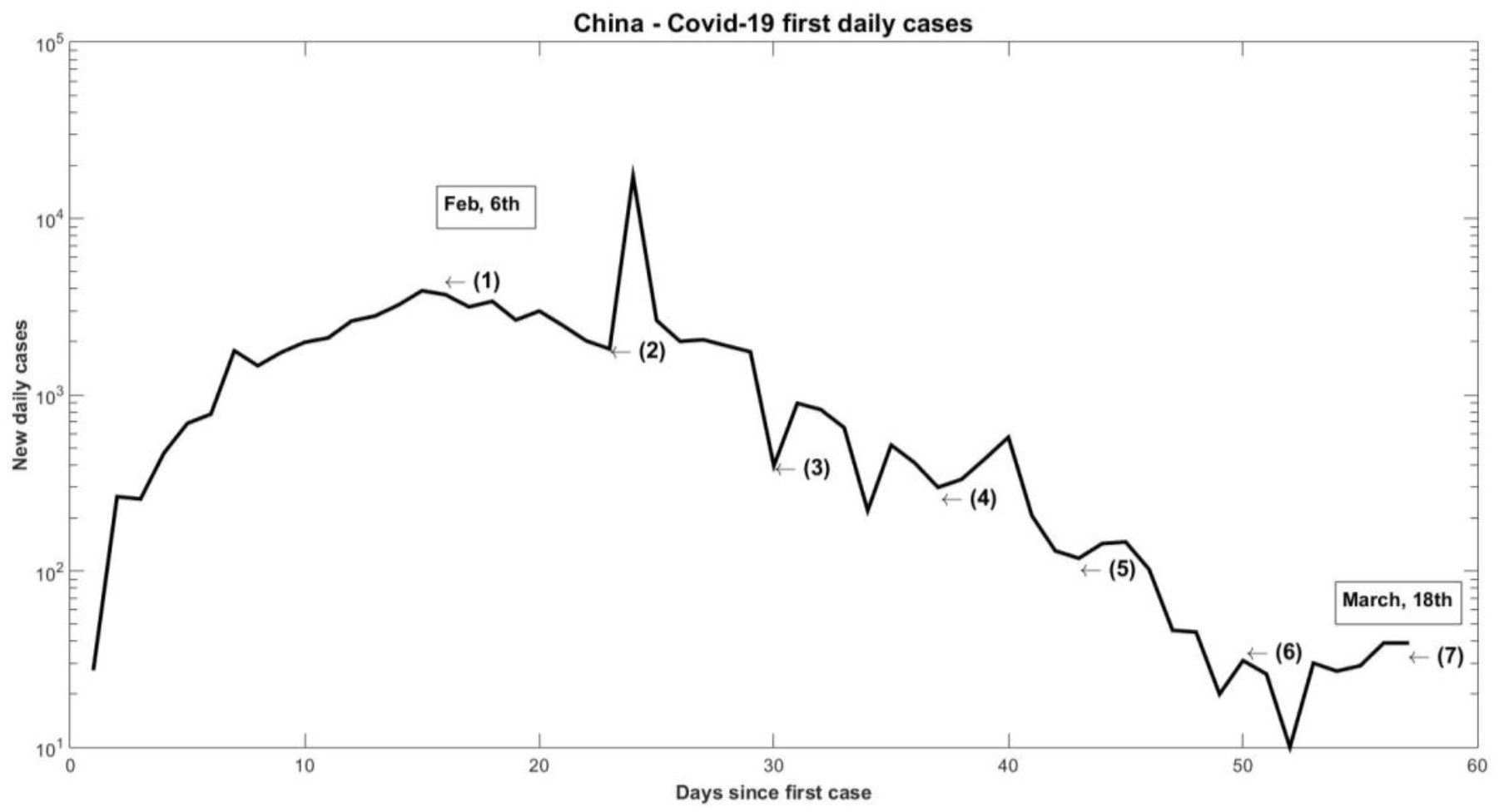
Daily new cases in China (February 2020)

From point (4) (March 23) the daily cases begin to recede, with the parameters moving in the direction of the bifurcation plane outside the bistable region. The large oscillation seen in Figure-18 between (4) and (8) is a reflection of the movement of the parameters back and forth in the bifurcation plane outside the region known as bistable. This great oscillation ends only as (κ,*r*) they move further and further away from the bistable region towards the origin.

### F. China 2020

SARS-Cov-2 was first identified in Whuhan, in the province of Hubei in China on December 1, 2019. The rapid and worldwide expansion has prompted WHO to declare the outbreak as a pandemic. The government decreed a lockdown in Wuhan on January 23, 2020 in an attempt to contain the spread. Figure-20 shows the new daily cases in China, observing a peak on February 14, jumping from 1,820 cases to 17,382 cases. Despite being in the offspring of contamination, this number was the result of a recount and review of different methodologies used at the time by health authorities in China.

Point (1) in Figure-20 notes the beginning of the decrease in the number of positive cases reported to WHO. Seven different points were noted in these data to observe the movement of the parameters (κ,*r*) in the bifurcation plane. The seven points are highlighted in Table-7 with their respective dates and number of reported cases.

**Table-7.** Covid-19 (κ,*r*) parameters identified for China.

|  | *Day* | *k* | *r* | *New cases* |
| --- | --- | --- | --- | --- |
| (1) | February, 6 | 84.34 | 0.17 | 3,697 |
| (2) | February, 13 | 1.92 | 0.34 | 1,820 |
| (3) | February, 20 | 7.06 | 0.18 | 395 |
| (4) | February, 27 | 8.33 | 1.02 | 298 |
| (5) | March, 4 | 6.9 | 3.03 | 118 |
| (6) | March, 11 | 5.72 | 0.44 | 31 |
| (7) | March, 18 | 17.9 | 0.17 | 39 |

**Table-8.**
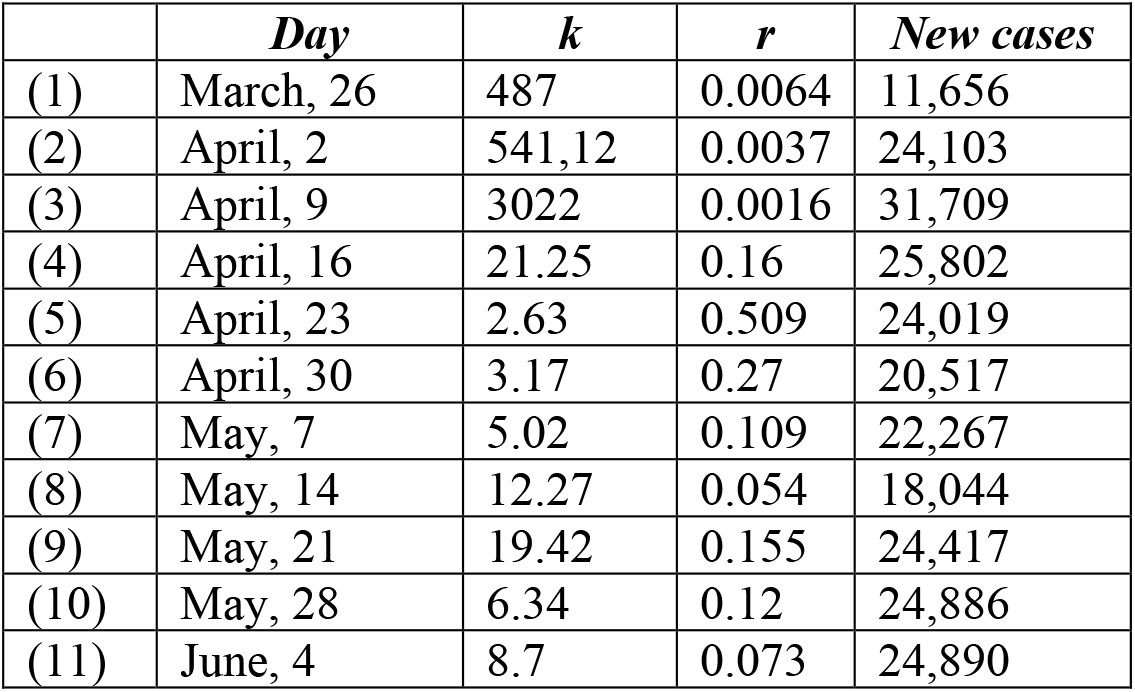
Covid-19 parameters (κ,*r*) identified for the USA.

|  | *Day* | *k* | *r* | *New cases* |
| --- | --- | --- | --- | --- |
| (1) | March, 26 | 487 | 0.0064 | 11,656 |
| (2) | April, 2 | 541,12 | 0.0037 | 24,103 |
| (3) | April, 9 | 3022 | 0.0016 | 31,709 |
| (4) | April, 16 | 21.25 | 0.16 | 25,802 |
| (5) | April, 23 | 2.63 | 0.509 | 24,019 |
| (6) | April, 30 | 3.17 | 0.27 | 20,517 |
| (7) | May, 7 | 5.02 | 0.109 | 22,267 |
| (8) | May, 14 | 12.27 | 0.054 | 18,044 |
| (9) | May, 21 | 19.42 | 0.155 | 24,417 |
| (10) | May, 28 | 6.34 | 0.12 | 24,886 |
| (11) | June, 4 | 8.7 | 0.073 | 24,890 |

For point (1) on February 6, it was calculated and observed that (κ,*r*) was within the bistable region on February 6. Figure-21 shows the configuration of the points, where it can be seen that when (κ, r) was in (1) the number of new cases in China on February 6 was 3,697 and for point (2), outside the bistable region the reduction was abrupt decreasing 50% in seven days. This confirms the downward trajectory that existed in China, in Figure-20, when the peak of cases reported and reviewed after the point (2) occurs.

Points (3) to (7) show the controlled outbreak in China, with the movement (κ,*r*) located in the refuge region in (3) and jumping into the outbreak region (4), however not passing by the bistable region. The movement from point (3) to point (7) was outside the bistable region, because although the rate of contagion increased, the carrying capacity was stable at the same value as the previous date.

If (κ,*r*) had entered the bistable region, we could have an even greater drop in the number of cases, or the return of a sudden increase. Everything depends on whether, when entering the bistable region, the parameters pass through the unstable point located in that region, or through one of the two existing stable points, predicted by the Catastrophe Theory.

### G. USA 2020

As in all countries, Covid-19 expanded rapidly in the USA with the first lockdowns being declared as of March 15, 2020. The first quarantines took place in Puerto Rico and the western USA. On March 20, it was New York’s turn to declare a lockdown with the threat of a lack of hospital beds in the city. After March 23, more states adhered to the policy of social distance.

Figure-22 notes as a point (1) March 26, 2020 when 11,656 new cases were reported in the USA. On that day the number of accumulated cases was 63,570 with 884 deaths reported to WHO. It can be seen in the figure the rapid expansion in the number of new cases up to point (2), on April 2, with a 106% increase in reported positive tests. And right after that, even with lockdown, point (3) shows a new increase of 31.5% a week later, on April 9th. Despite the fact that point (3) shows a smaller increase in the number of cases, the fatality of the disease increased, since the number of deaths jumped from 884 to 12,749 in just two weeks.

**Figure-21.**
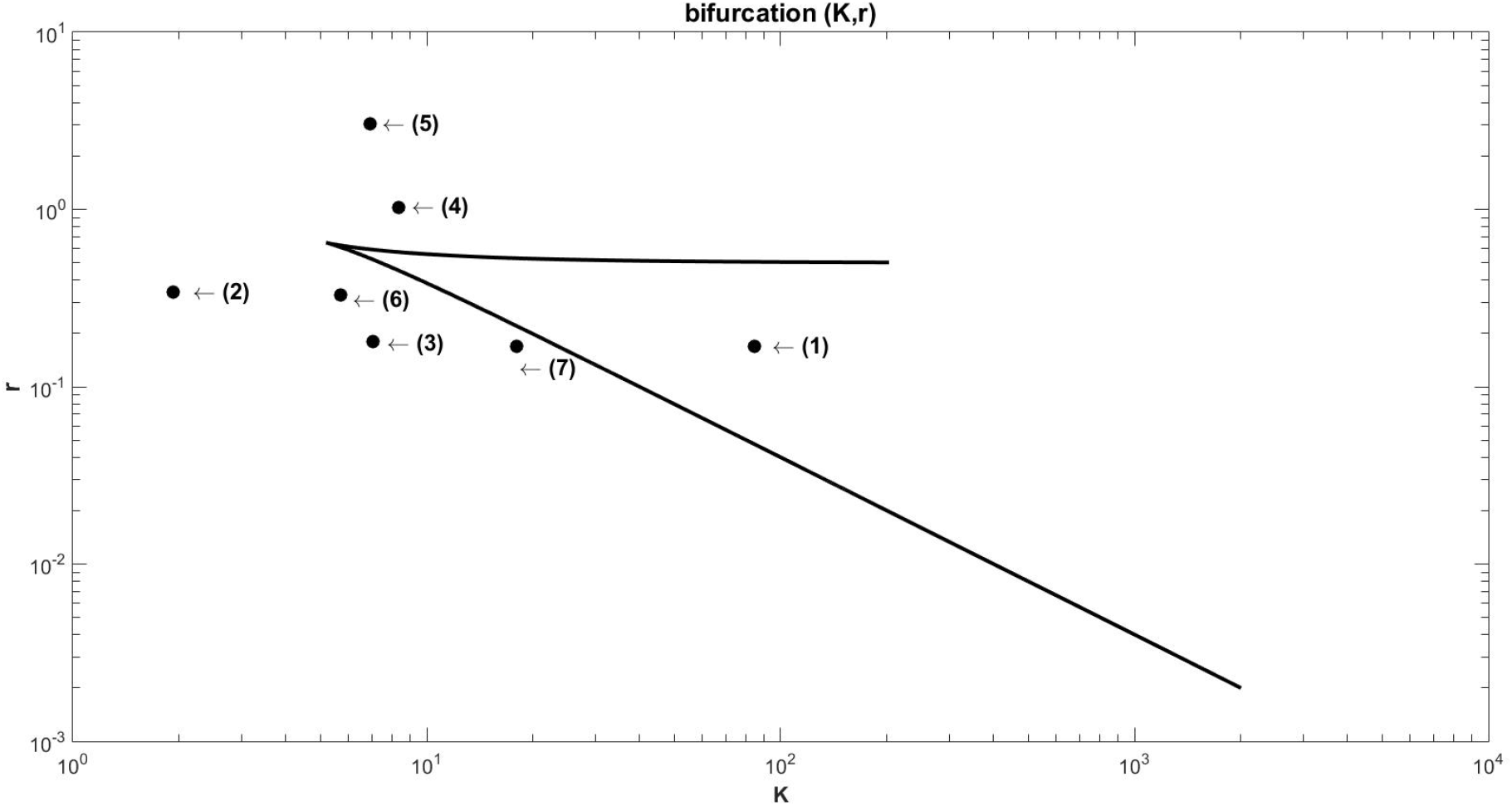
Bifurcation plan with estimated parameters (κ,*r*) for China.

**Figure-22.**
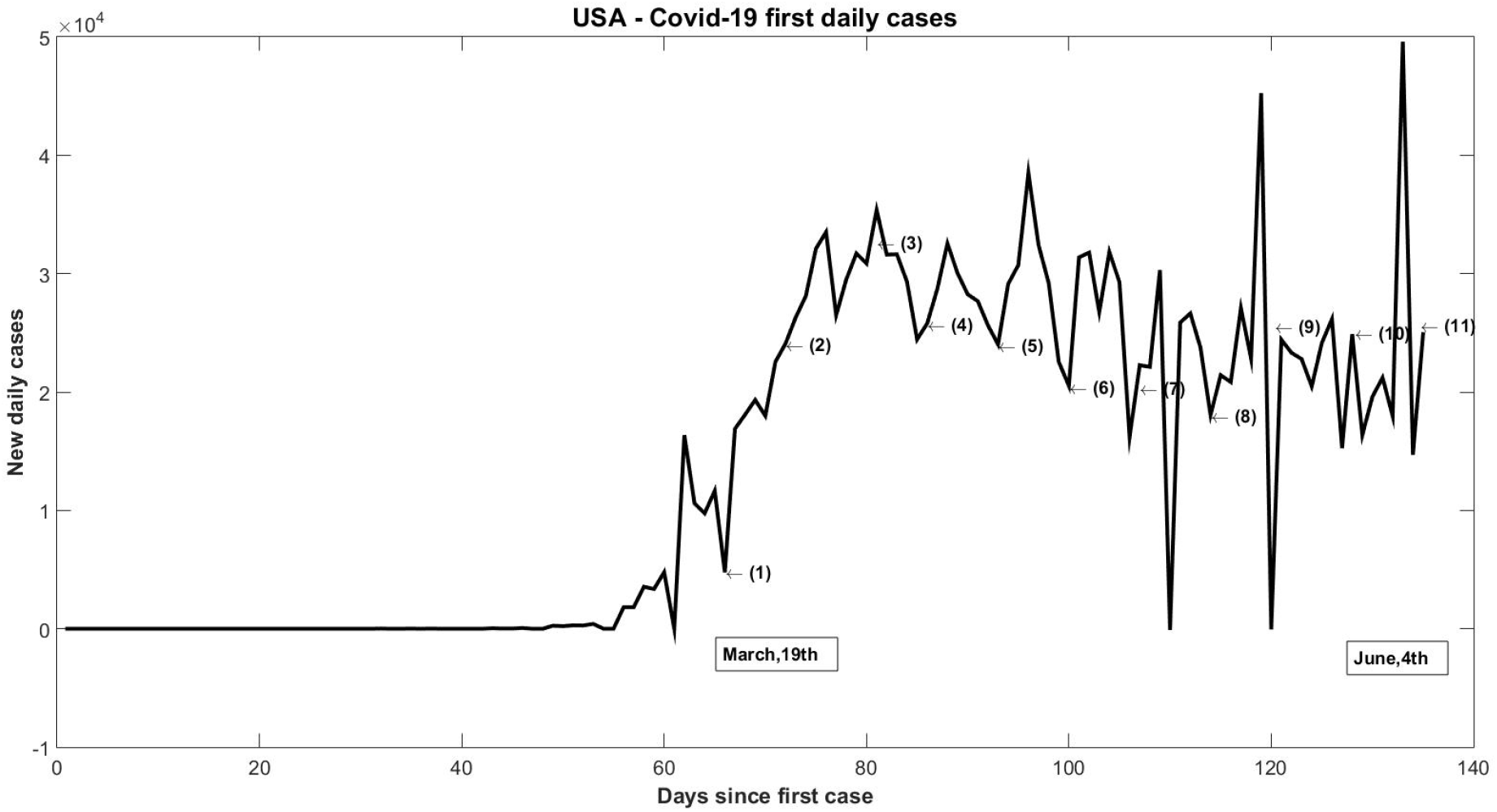
New Daily Cases in the USA (March - June 2020)

**Figure-23.**
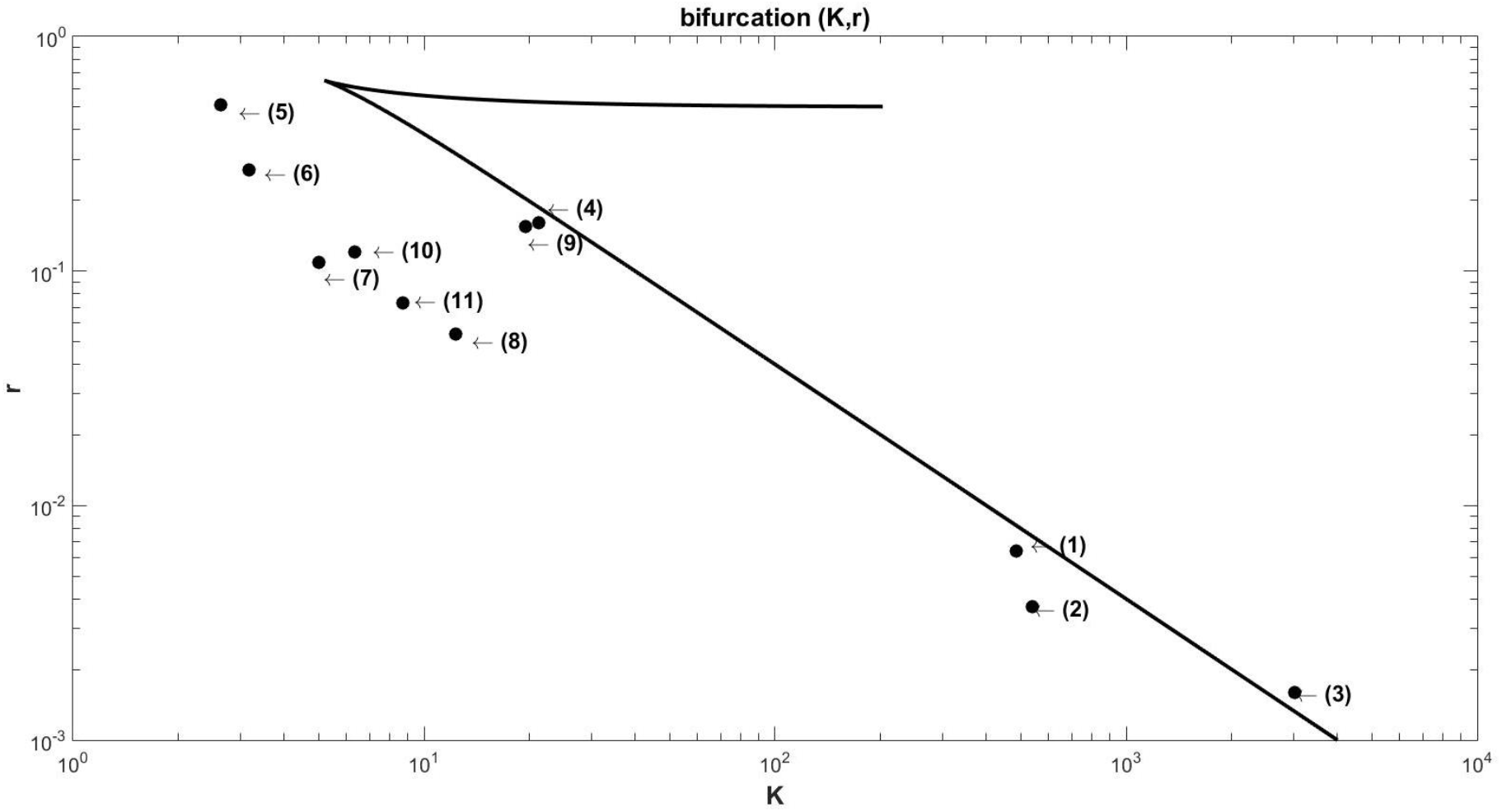
Bifurcation plan with estimated parameters (κ,*r*) for the USA.

Figure-23 represents points (1) to (11) in Table-8 compared to the same data as Figure-22. Following the parameter identification algorithm (κ,*r*), the movement in the bifurcation plane can be observed over the weeks. On the most severe days of the disease between March 26 and April 9, (κ,*r*) were close to the bistability region, jumping into that region in (3). The increase of more than 100% in new cases from point (1) to point (3) exactly coincides with the entry of (κ,*r*) into the interior of the bistable region, an abrupt jump in the most acute period of the pandemic in the USA.

When (κ, r) left the bistable region in (4) the cases decreased slightly to 25,802 positive cases daily. Since then, from (4) to (11) the number of daily positive cases has become stable, but very high. The average number of cases in this period was 25,462 and it can be seen in Figure-23 that (κ,*r*) does not converge to the region known as a refuge in the bifurcation plane, where the points are always close to the limit of the bistable region. From the point of view of Figure-22, this location causes a great oscillation around 25 thousand cases in points (4) to (11) and does not decrease consistently.

In this region of Figure-23, the carrying capacity is stable with the reproduction rate r high and fluctuating with each new day. In order for the number of positive cases to decrease consistently, it was necessary for r to converge towards zero and for the support capacity *k* to decrease close to the origin in the bifurcation plane. An uncontrolled reopening can take the parameters (κ,*r*) back to the bistable region and cause a new explosion of cases.

## IV. DISCUSSION

Since the beginning of the data released by WHO on Covid-19, we observed that positive cases are stable and few in the beginning of the disease. And then, abruptly, an explosion of contamination occurs, forcing all countries to follow a severe public policy of social restriction.

Positive cases from countries that controlled the pandemic, show through the data that the decrease in contamination comes after controlling the great daily fluctuation in the case reports. When these oscillations decrease their range of variability, the accumulated contamination curve begins to converge while the daily contamination curve begins its fall period.

It was highlighted in the cases presented for the countries Italy, South Korea, Brazil, United Kingdom, France, China and the USA that the same behavior in the oscillation of the data and the sudden explosion of the cases for the contamination by SARS-Cov-2 take the non-dimensional parameters (κ, r) into a bistable region in the bifurcation plane. The sudden entry or exit of this region presents an explosion or a substantial and sudden decrease in the number of positive cases for Covid-19.

According to the Catastrophe Theory, this exit to the lower region and converging to the origin indicates refuge from contagion, identifying that the outbreak of the disease is under control. Likewise, leaving the bistable region to a higher region would be identified as being out of control in the fight against the pandemic. The simple passage through the bistable region can lead the number of contagions to the explosion status of cases, as on the contrary, a sudden and abrupt decrease.

In some cases, this representation was quite plausible and coincided with daily positive case data. In the Italian data, when (κ,*r*) left the bistable region, the sudden increase from one week to the next was 54.8%, after which the data remained converging slowly towards the refuge region. As (κ,*r*) did not pass through the bistable region on its return to the refuge, the drop in the number of daily cases in Italy was slow and with great oscillation.

In the case of Korea, we took data after the critical phase of the pandemic to verify the behavior of (κ,*r*) when Covid-19 was in its control phase. The oscillation of the data is much less since the passage outside the bistable region was fast, with low expansion value r despite high carrying capacity k in the last collected data. It can be seen that in Korea, already in the control phase of Covid-19, there was a significant drop with the passage and exit of (κ,*r*) through the bistable region.

In the case of Brazil, there was a large fluctuation in the daily positive case data, with two internal points in the bistable region. The oscillation in the cases is high and always increasing for the observed period. In the case of UK two points were within the bistable region, exactly coinciding with the period of greatest expansion of Covid-19. With the sampled data, when (κ,*r*) left the region, it remained very far from the origin known as a refuge in the Catastrophe Theory. Despite the low expansion rate, the support capacity was still high in this sample, revealing stability in the contagion curve, but at a high level and with great oscillation in the cases (between 4 thousand and 6 thousand positive daily cases).

Before the explosion of cases in France, (κ,*r*) presented 3 points within the bistable region and when it left the region the number of cases increased suddenly. After the increase, (κ,*r*) left the bistable region, but just like the UK, it moved away from the region of refuge, also showing a strong fluctuation in the number of daily cases. Despite presenting a drop in the daily contagion curve, the oscillation of cases was still around 3 thousand a day in this period.

In the case of China (κ, r) it passed through the bistable region only once, where on leaving it coincides with a sharp drop in the number of daily cases. No new passage through the bistable region was detected, but this may have occurred quickly and in a shorter period than the sample. This is because two consecutive points appear above and below the region, with a substantial drop in the number of daily cases. It was not possible to observe this passage, because for very small differences in the number of accumulated cases, the identification of (κ,*r*) using the steps of the presented algorithm is not accurate.

For the USA data, the passage of (κ, r) through the bistable region coincided with an increase of more than 100% in positive cases in the country. The jump was sudden in terms of both positive cases and deaths in just two weeks. After (κ, r) left the bistability region, the number of daily cases stabilized, but at a very high level. It is possible to observe in the plane of bifurcation of parameters that both the expansion r and the carrying capacity k oscillated outside the bistable region but in a circle. It can be seen that when r decreases, k increases, and at other points, r increases when k decreases. Unlike China, which in the last few points has decreased, in the USA data there is an oscillation of this parameter. This may reflect the strong fluctuation in the final data for the sampling of positive cases in the USA.

## V. CONCLUSION

Catastrophe Theory and Bifurcation have very wide approaches and frequent applications in several areas of Science. In this work, the use of the bifurcation methodology for outbreaks was presented, especially for insect outbreaks (Ludwig, 1978), adapting the approach to SARS-Cov-2 in the Covid-19 pandemic. With data obtained from WHO for 7 countries, an algorithm was presented to identify the rate of expansion of the disease and the carrying capacity.

To test the algorithm, a first single example was used with Spanish flu data between 1918-1920, using the particle swarm algorithm to identify the parameters of the logistical curve that represents the number of accumulated cases (or accumulated deaths). With these parameters identified and using the necessary transformations to make the model variables non-dimensional. On a case-by-case basis, for each different country, we sought to verify positions of passage of these parameters through the region of a “cusp” curve predicted by the Catastrophe Theory also used in the theory of bifurcation.

In these theories, the bifurcation plan presents three distinct regions: refuge, bistable and outbreak. Using the same notion to detect the movement of parameters in the case of an insect outbreak, new parameters were identified for each country and each week, cataloging their movements and comparing the number of positive cases for SARS-Cov-2.

In all countries studied with these data collected from WHO, it was observed that before a strong and controlled fall, strong fluctuations occur in these data. It can also be observed that before stabilization and after weeks of few isolated cases, sudden and abrupt increases occur in the reported cases. The sudden increase in cases was coincident in the data collected from countries with the passage of the expansion rate r and carrying capacity K through the region of the Catastrophe Theory known as bistable. When leaving this region, depending on whether it is at the top or bottom, an abrupt increase or decrease in cases is observed in the data.

There is much discussion in all countries about the reopening of trade, the reopening of the economy for the post-pandemic period. In addition to the traditional R0 or Rt parameter of epidemiology, used as a measure of the spread of contamination, the observation of the parameters (κ,*r*) and their movement towards the bistable region, can contribute to a further understanding of the exact moment of an outbreak and whether there is indeed a control on contamination. For instance, if Rt is growing and parameters (κ, r) are in bistable region, is possible that a new outbreak will begin (or second wave of contamination).

In the outbreak control period, when the number of accumulated cases in a week is very close to the number of accumulated cases in the previous week, the identification is not very accurate. But for the most critical period of the expansion, where a safe interpretation is necessary, the parameterization through (κ,*r*) as well as its movement in the bifurcation plane are quite coincident and can predict problems, in addition to assisting in the containment strategy of the outbreak.

Number of positive controlled cases, but with high daily oscillation, reflect that the disease is outside the bistable region. But with few changes (relaxation of the quarantine, not isolation) they can bring the parameters back into that region (κ,*r*). And if that occurs, a new period of expansion of the outbreak can be observed. The function p(N) is a very important representation of actuation of social policy, indicating that high values decreasing outbreak of Covid-19, showing success of control, with positive results for social distancing. Is possible indicate that Catastrophe Theory can be used as a new alert of new outbreaks or new waves of expansion for SARS-Cov-2, but is highly necessary a good precision in data acquisition to fit correctly parameters of cusp curve.

## DATA AVAILABILITY

The data that support the findings of this study are available within the article.

## Data Availability

All data in this article were acquired directly from WHO.

https://covid19.who.int/table

## ACKNOWLEDGEMENTS

The author is deeply grateful to professor Dr. Marcus Werner Beims (Universidade Federal do Paraná – Brazil), for the valuate, revision and incentives for this article.

## REFERENCES

1. Abdeljaoued, I. A pandemic at the Tunisian scale Mathematical modelling of reported and unreported COVID-19 infected cases. 2020. medRxiv doi:https://doi.org/10.1101/2020.05.21.20108621 accessed in May/23/2020.

2. Abramov, D.M., Gomes Junior, S.C. Multivariate Prediction Network Model for epidemic progression to study the effects of lockdown and coverage on a closed community on theoretical and real scenarios of COVID-19. 2020. medRxiv doi:https://doi.org/10.1101/2020.05.04.20090712 accessed in May/22/2020.

3. Anderson, R. M., May, R. M. Infectious Diseases of Humans: Dynamics and Control. Oxford Univ. Press, 1992. 757p.

4. Anderson, R. M., May, R. M. Population biology of infectious diseases I. Nature 280, (1979), 361–367.

5. Anderson, R. M., May, R. M. Vaccination and herd immunity to infectious diseases. Nature 318, (1985), 323–329. Arnold, V. I. Catastrophe Theory. Springer-Verlag, New York. (1986), 108 p.

6. Bjørnstad, O.N., Shea, K., Krzywinski, M., Krzywinski, M., Altman, N. Modeling infectious epidemics. Nat Methods 17, 455–456 (2020).https://doi.org/10.1038/s41592-020-0822-z

7. Casti. J. Catastrophes, control and the inevitability of spruce budworm outbreaks. Ecol. Modelling, 1982, (14) 293–300.

8. Castrigiano, D. P. L., Hayes, S. A. Catastrophe Theory. CRC Press Taylor & Francis Group, Boca Raton FL – USA. (2004), 264 p.

9. Chintalapudi, N., Battineni, G., Amenta, F. COVID-19 virus outbreak forecasting of registered and recovered cases after sixty day lockdown in Italy: A data driven model approach. Journal of Microbiology, Immunology and Infection 2020, 1–8.https://doi.org/10.1016/j.jmii.2020.04.004

10. Chitanvis, S. M. Dynamical model for social distancing in the U.S. during the COVID-19 epidemic. 2020. medRxiv doi:https://doi.org/10.1101/2020.05.18.20105411 accessed in May/22/2020.

11. Collins, S., Frost, W.H., Gover, M., Sydenstricker, E. Mortality from Influenza and Pneumonia in 50 Large Cities of the United States, 1910-1929. Public Health Reports (1896 – 1970). Association of Schools of Public Health. Vol. 45, No. 39 (Sep. 26, 1930), 2277–2328.http://www.jstor.org/stable/4579795

12. Cotta, R.M., Naveira-Cotta, C.P., Magal,P. Parametric identification and public health measures influence on the COVID-19 epidemic evolution in Brazil. 2020. medRxiv doi:https://doi.org/10.1101/2020.03.31.20049130 accessed in May/22/2020.

13. Cuevas, E. An agent-based model to evaluate the COVID-19 transmission risks in facilities. Computers in Biology and Medicine 2020, (121) 1–12 https://doi.org/10.1016/j.compbiomed.2020.103827

14. Gilmore, R. Catastrophe Theory for Scientists and Engineers. Dover. (1993), 666 p.

15. Giordano, G., Blanchini, F., Bruno, R., Colaneri, P., Di Filippo, A., Di Matteo, A., Colaneri, M. Modelling the COVID-19 epidemic and implementation of population-wide interventions in Italy. Nat Med (2020).https://doi.org/10.1038/s41591-020-0883-7

16. Ivorra, B., Ferrández, M.R., Vela-Pérez, M., Ramos, A.M.Mathematical modeling of the spread of the coronavirus disease 2019 (COVID-19) taking into account the undetected infections. The case of China. Commun Nonlinear Sci Numer Simulat 2020, (88) 1–4 https://doi.org/10.1016/j.cnsns.2020.105303

17. John Hopkins University & Medicine. Coronavirus Resource Center. 2020. Disponível em :https://coronavirus.jhu.edu/map.html acessado em 22 de Maio 2020.

18. Kang, D., Choi, H., Kim, J.H, Choi, J. Spatial epidemic dynamics of the COVID-19 outbreak in China. International Journal of Infectious Diseases 2020, (94) 96–102. https://doi.org/10.1016/j.ijid.2020.03.076

19. Kucharski A.J., Russell, T.W., Diamond, C., Liu, Y., Edmunds, J., Funk, S., Eggo, R.M. Early dynamics of transmission and control of COVID-19: a mathematical modelling study. Lancet Infec Dis 2020. (20) 553–558.

20. Liang, K. Mathematical model of infection kinetics and its analysis for COVID-19,SARS and MERS. Infection, Genetics and Evolution 2020, (82) 1–7. https://doi.org/10.1016/j.meegid.2020.104306

21. Liu, L., Liu, W., Cartes, D. A. Particle swarm optimization-based parameter identification applied to permanent magnet synchronous motors. Engineering Applications of Artificial Intelligence. 2008. (21) 1092–1100.https://doi.org/10.1016/j.engappai.2007.10.002

22. Liu, Z., Magal, P., Seydi, O., Webb, G. A COVID-19 epidemic model with latency period. Infectious Disease Modelling 2020, (5) 323–337.https://doi.org/10.1016/j.idm.2020.03.003

23. Ludwig, D., Jones, D. D., Holling, C. S. Qualitative Analysis of Insect Outbreak Systems: The Spruce Budworm and Forest. The Journal of Animal Ecology, Vol. 47, No. 1. 1978. 315–332.

24. May, R., Anderson, R. Transmission dynamics of HIV infection. Nature 326, 137–142 (1987).https://doi.org/10.1038/326137a0

25. Mlocek, W., Lew, R. Forecasting Trajectories of an Emerging Epidemic with Mathematical Modeling in an Online Dashboard: the Case of COVID-19. 2020. medRxiv doi:https://doi.org/10.1101/2020.05.21.20108753 accessed in May/23/2020.

26. Ndairou, F., Area, I., Nieto, J.J., Torres, D.F.M. Mathematical Modeling of COVID-19 Transmission Dynamics with a Case Study of Wuhan. Chaos, Solitons and Fractals 2020, 1–11, doi:https://doi.org/10.1016/j.chaos.2020.109846

27. Neyens, T., Faesa, C., Vranckxa, M., Pepermansc, K., Hensa, N., Van Dammed, P., Molenberghsa, G., Aertsa, J., Beutels, P. A spatial model to optimise predictions of COVID-19 incidence risk in Belgium using symptoms as reported in a large-scale online survey. 2020. medRxiv doi:https://doi.org/10.1101/2020.05.18.20105627 accessed in May/22/2020.

28. Nowak, M. A., Anderson, R. M., Mclean A. R., Wolfs T. F., Goudsmit, J. and May, R. M. Antigenic Diversity Thresholds and the Development of AIDS. Science 254 (1991). 963 – 969.

29. Nowak, M. A., May, R. M, Phillips R. E., Jones, S. R., Lallo, D. G., McAdams, S., Klenerman, P., Köppe, B., Sigmund, K., Bangham, C. R. M. and McMichael, A. J. Antigenic oscillations and shifting immunodominance in HIV-1 infections. Nature 375 (1995). 606–611.

30. Postnikov, E. B. Estimation of COVID-19 dynamics “on a back-of-envelope”: Does the simplest SIR model provide quantitative parameters and predictions? Chaos, Solitons and Fractals 2020, (135) 1–6. https://doi.org/10.1016/j.chaos.2020.109841

31. Robeva, R., Murrugarrab, D. The spruce budworm and forest: a qualitative comparison of ODE and Boolean models. Letter in Biomathematics, Vol 3, no.1, 2016, 75–92. http://dx.doi.org/10.1080/23737867.2016.1197804

32. Sagar, A., LeCover, R., Shoemaker, C., Varner, J. Dynamic Optimization with Particle Swarms (DOPS): A meta-heuristic for parameter estimation in biochemical models. (2017). 1–30. bioRxiv doi:https://doi.org/10.1101/240580

33. Sendrescu, D. and Roman, M. Parameter identification of bacterial growth bioprocesses using particle swarm optimization. 2013 9th Asian Control Conference (ASCC), Istanbul, 2013, pp. 1–6, doi:10.1109/ASCC.2013.6606279.

34. Woodcock, A., Davis, M. Catastrophe Theory. E. P. Dutton, New York. (1978), 152 p.

35. World Health Organization(^a^) – WHO Coronavirus Disease (COVID-19). 2020. Available in :https://covid19.who.int/ accessed in May/22/2020.

36. World Health Organization(^b^) - WHO Coronavirus disease 2019 (COVID-19). Situation report 24.February 13, 2020. Geneva: World Health Organization, 2020.

37. Xie, Y., Fu, J., Chen, B. Parameter identification of hysteresis nonlinear dynamic model for piezoelectric positioning system based on the improved particle swarm optimization method. Advances in Mechanical Engineering. 2017, 9(6) 1–12. DOI: 10.1177/1687814017702813

38. Yang, H. M., Lombardi Junior, L.P., Castro, F.F.M., Yang, A.C. Evaluating epidemiological scenarios of isolation and further releases considering protection actions to control transmission of CoViD-19 in São Paulo State, Brazil. medRxiv doi:https://doi.org/10.1101/2020.05.19.20099309 accessed in May/22/2020.

39. Yousefpour, A., Jahanshahi, H., Bekiros, S. Optimal policies for control of the novel coronavirus disease (COVID-19) outbreak. Chaos, Solitons and Fractals 2020, (136). https://doi.org/10.1016/j.chaos.2020.109883

40. Zeeman, E. C. Catastrophe Theory-Selected Papers 1972 - 1977.Addison-Wesley,(1977), 675 p.

